# From Anatomy to Aneurysm: Morphological-Hemodynamic Coupling and Predictive Modeling in Aberrant Splenic Artery

**DOI:** 10.64898/2026.09.08.26362579

**Authors:** Ran Xu, Shouji Qiu, Tianyue Pan, Zhaohui Hua, Ding Yuan, Xiaolong Wei, Meng Ye, Yaoguo Yang, Minyi Yin, Ye Tian, Qingbo Fang, Youfei Qi, Donghua Ji, Hongping Deng, Haotian Chi, Yuan Feng, Haozhe Qi, Tiehao Wang, Li Chen, Mojia Hu, Wenzhen Yang, Yuqi Yi, Ruchen Li, Tao Lyu, Wei Wang, Donglin Li, Lixin Wang

## Abstract

**Background:** Aberrant splenic artery (SA) is an extremely rare anatomical variant in which the SA arises from the superior mesenteric artery (SMA) rather than the celiac axis. Individuals with this anatomical variant are at markedly increased risk of developing SA aneurysms (SAAs). However, no systematic investigation into its underlying pathogenesis has been conducted to date, nor has a pathogenesis-based predictive model been developed to assess future aneurysm risk in this population.

**Objectives:** This study aimed to systematically investigate the morphological and hemodynamic mechanisms by which aberrant SA anatomy predisposes to local SAA formation, and to develop an interpretable predictive model for individualized risk stratification based on these mechanisms.

**Methods:** This multicenter retrospective cohort study enrolled 195 patients with aberrant SA from 16 centers across China between January 2008 and June 2026, including 94 with concomitant aberrant SAAs and 101 without. 3D vascular models were reconstructed from CTA images, and 12 morphological parameters were measured. Computational fluid dynamics simulations were performed to analyze 14 hemodynamic parameters. Ultimately, 5 morphological parameters were selected to construct a two-stage interpretable machine learning model for predicting the occurrence and location of SAAs.

**Results:** Morphologically, aberrant SAAs were significantly larger (diameter: 20.4mm vs. 17.5mm, *P*<0.001; length: 20.5mm vs. 17.9mm, *P*=0.002) and predominantly located in the proximal SA, whereas common SAAs were mainly located in the distal splenic hilum. Furthermore, the aberrant SA exhibited a smaller branching angle with its parent vessel (22.3° vs. 19.7°, *P*=0.010), an increased SA-to-SMA cross-sectional area ratio (0.57 vs. 0.47, *P*=0.008), and a higher tortuosity index (*P*<0.001). Hemodynamic analysis demonstrated significantly lower TAWSS at the proximal SA in aberrant anatomy (*P*<0.001), whereas the OSI at the SA-parent vessel junction was significantly elevated (*P*=0.004) and inversely correlated with the branching angle (*P*<0.001, r=-0.686). The interpretable machine learning model, built on morphological and hemodynamic principles, achieved an AUROC of 0.828 for predicting aberrant SAA occurrence and was deployed as an interactive web application (the zs_abeSA model) for clinical use.

**Conclusions:** Aberrant SA anatomy is an independent risk factor for SAA formation. The pathogenic mechanism involves a cascade from morphological remodeling to hemodynamic derangement, ultimately leading to aneurysm formation at the proximal SA. The zs_abeSA model provides a practical tool for individualized risk assessment, with direct implications for early screening and surveillance strategy development.

## Introduction

Visceral artery aneurysms (VAAs) are a rare disease with an overall incidence ranging from 0.1% to 2% [1]. Approximately 25% of patients with VAAs present emergently due to aneurysm rupture, with an associated mortality rate of 8.5% [2]. Splenic artery aneurysms (SAAs) are the most common subtype of VAAs, accounting for 60% of all VAAs, and are also the third most common abdominal aneurysms after aortic and iliac artery aneurysms [3,4]. Among them, SAAs resulting from the congenital anatomical variation in which the splenic artery (SA) arises from the superior mesenteric artery (SMA), also referred to as aberrant SAAs, represent a rare but distinct subtype with a low incidence. Nevertheless, these aneurysms warrant close clinical attention and early intervention, given that their potential risk of rupture may lead to catastrophic outcomes. Since the first case of this type of SAA was reported by Ghatan *et al.* in 1967, comprehensive and systematic investigations into its pathogenesis have remained scarce, likely due to the limited number of cases, a lack of diverse research methodologies, and technological constraints at the time [5]. Therefore, there is an urgent need to integrate large-scale clinical data with novel research methodologies to elucidate the pathogenic mechanisms and enable individualized risk assessment and surveillance planning for these patients.

Recently, image-based computational fluid dynamics (CFD) models have emerged as a powerful tool for investigating various cardiovascular diseases [6]. Numerous hemodynamic parameters have been demonstrated to be associated with the growth rate or rupture risk of cerebral or aortic aneurysms, including wall shear stress (WSS), oscillatory shear index (OSI), gradient oscillatory number (GON), and aneurysm formation index (AFI). These parameters can be determined noninvasively using imaging and computational modeling, making them potential biomarkers for future risk intervention and management of aneurysmal disease [7]. However, owing to the low incidence of VAAs (particularly SAAs), systematic hemodynamic investigations using CFD have long been lacking.

Machine learning (ML) has emerged as a transformative technology in vascular disease research, opening new avenues for predictive modeling, risk stratification, and clinical decision support [8]. For abdominal aortic aneurysms, ML models have demonstrated considerable potential in predicting aneurysm growth and rupture risk. For instance, geometric ML models have been applied to directly predict localized AAA growth on three-dimensional vascular surfaces, achieving high accuracy in identifying patients suitable for surgical repair [9]. Similarly, XGBoost and other ML algorithms have been used to stratify rupture risk based on parameters such as vascular tortuosity, intraluminal thrombus thickness, and wall stress [10]. In VAAs, AI-based methods have been explored for automated detection from CTA images, achieving a sensitivity of 0.93 in identifying VAAs [11]. However, the application of ML to SAAs remains limited. Previous studies have primarily relied on traditional statistical methods, such as Cox proportional hazards modeling, to identify predictors of SAA expansion, including initial diameter >14mm, smoking history, and absence of wall calcification [12]. To date, no ML-based predictive model has been specifically developed for SAAs, particularly in the context of aberrant SA anatomy.

In this study, we systematically collected clinical data and imaging-based anatomical characteristics from a multicenter cohort, and constructed a CTA-based CFD model to analyze their aberrant morphological and hemodynamic features. Based on these data, we developed an interpretable predictive model to assess the risk of aneurysm occurrence and its potential location in patients with aberrant SA anatomy, which has the potential to substantially optimize the current diagnostic, therapeutic, and surveillance workflow for this rare disease.

## Methods

### Study population

This study was approved by the Committee for the Protection of Human Subjects at Zhongshan Hospital, Fudan University, as well as the institutional review boards of all participating centers, and informed consent was obtained from all enrolled patients. This multicenter retrospective study included patients with SAAs admitted to 16 centers across China between January 2008 and June 2026. The patient screening process at each center is summarized in **Figure 1A** and **Table 1**. Briefly, by querying each center‘s computed tomography angiography (CTA) database, a total of 1,197 patients with anatomical anomalies of the SMA were preliminarily identified from 25,770 patients diagnosed with SA anomalies or pathology. After further excluding patients with a history of trauma or surgery involving the SMA or SA region (n=179), those with the proper hepatic artery arising from the SMA (n=714), and those with poor image quality (n=109), 195 patients with definitive aberrant SA anatomy (i.e., the SA arising from the SMA) were finally included for this study. Among them, 94 patients developed SAAs in the setting of this aberrant anatomy, while 101 did not.

**Figure 1.**
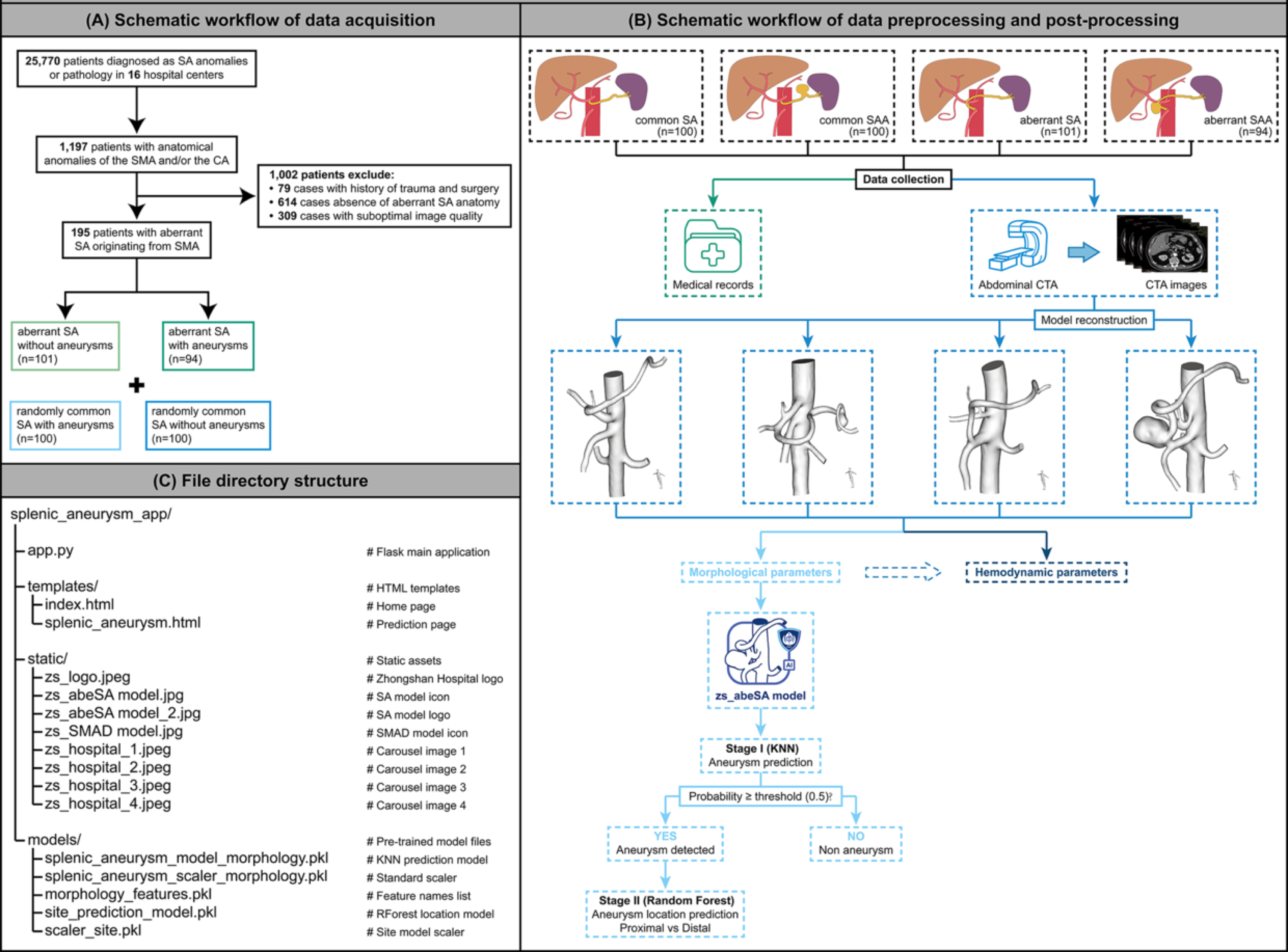
Study flowchart and architecture of the zs_abeSA model. **(A)** Patient screening flowchart. From 25,770 patients diagnosed with SAAs, 1,197 cases with SMA anatomical anomalies were identified through CTA database screening. After excluding those with a history of trauma or surgery (n=79), those with the proper hepatic artery arising from the SMA (n=614), and those with poor image quality (n=309), 195 patients with definitive aberrant SA anatomy were ultimately enrolled, including 94 with concomitant SAAs and 101 without. **(B)** Overview of the overall study design. This includes morphological measurements, CFD analysis, ML modeling, and the development of the zs_abeSA model across the four study groups. **(C)** Schematic diagram of the zs_abeSA model web application architecture, adopting a client-server model. After users input five morphological parameters, the system sequentially calls the occurrence prediction model (KNN) and the location prediction model (random forest), returning the occurrence probability, risk level, and predicted location. CFD, computational fluid dynamics. CTA, computed tomography angiography. KNN, K-Nearest Neighbors. SA, splenic artery. SAA, splenic artery aneurysm. SMA, superior mesenteric artery.

**Table 1.** Flowchart of enrolled patients. CA, celiac axis. CHA, common hepatic artery. SA, splenic artery. SMA, superior mesenteric artery.

| Table 1. Flowchart of enrolled patients |  |  |  |  |  |  |  |  |  |  |  |  |  |  |  |  |
| --- | --- | --- | --- | --- | --- | --- | --- | --- | --- | --- | --- | --- | --- | --- | --- | --- |
| Centers |  |  |  |  |  |  |  |  |  |  |  |  |  |  |  |  |
| Number of diagnosed SA anomalies or pathology patients | No.1<br>6,339 | No.2<br>431 | No.3<br>637 | No.4<br>1,056 | No.5<br>930 | No.6<br>7,550 | No.7<br>3,821 | No.8<br>442 | No.9<br>670 | No.10<br>437 | No.11<br>454 | No.12<br>812 | No.13<br>113 | No.14<br>290 | No.15<br>377 | No.16<br>1,411 |
| CTA reports indicated anatomical anomalies of the SMA and/or the CA |  |  |  |  |  |  |  |  |  |  |  |  |  |  |  |  |
| Number of SA patients eligible for inclusion | 264 | 22 | 27 | 53 | 37 | 395 | 175 | 21 | 28 | 22 | 19 | 32 | 7 | 13 | 16 | 66 |
| • History of trauma and surgery | 36 | 5 | 7 | 4 | 5 | 21 | 14 | 3 | 4 | 3 | 1 | 7 | 2 | 3 | 5 | 11 |
| • Absence of aberrant SA anatomy | 142 | 11 | 15 | 27 | 14 | 214 | 106 | 15 | 14 | 15 | 9 | 19 | 1 | 8 | 9 | 43 |
| • Suboptimal image quality | 34 | 3 | 0 | 15 | 10 | 106 | 26 | 0 | 5 | 1 | 0 | 0 | 0 | 0 | 0 | 9 |
| Number of patients included in this study | 52 | 3 | 5 | 7 | 8 | 54 | 29 | 3 | 5 | 3 | 9 | 6 | 4 | 2 | 2 | 3 |

To establish reference values for physiological WSS, control cases were matched to aberrant SA patients based on a ‘±3 months’ criterion relative to the examination date. A cohort of patients with SAAs without anatomical anomalies (n=100) and a cohort of individuals without a history of abdominal aortic disease (n=100) were enrolled as control groups for hemodynamic analysis. Electronic medical records and follow-up information for all enrolled patients were independently extracted and reviewed by two investigators at each center. Documented data included patient demographic characteristics, treatment strategies, and perioperative outcomes. In addition, imaging data were collected to evaluate the morphological and hemodynamic characteristics of the aneurysms and the local vasculature.

### Study procedures

**Figure 1B** presents a schematic diagram of the data acquisition, preprocessing, and postprocessing workflow used in this study. The workflow comprised four main steps: (1) collection of clinical data and CTA images from the enrolled subjects; (2) extraction of morphological parameters by reconstructing and segmenting three-dimensional surface models of the abdominal aorta (ranging from approximately 10cm above the celiac axis (CA) bifurcation to approximately 10cm below the SMA bifurcation) and its branch arteries; (3) acquisition of hemodynamic parameters using computational fluid dynamics, including three-dimensional volumetric model construction, mesh generation, boundary condition setting, and hemodynamic parameter extraction; and (4) construction of a disease progression prediction model based on ML.

Two research assistants were trained to perform the above workflow under the guidance of an experienced radiologist and a CFD expert. Following this workflow, we obtained three-dimensional reconstruction models for four study groups (healthy control group, common SAA group, aberrant SA group, and aberrant SAA group) and extracted key morphological and hemodynamic parameters. A total of 12 morphological parameters closely associated with aneurysm formation and rupture, as well as 14 hemodynamic parameters from different vascular locations, were extracted.

### Clinical data collection

Medical records were reviewed by two experienced vascular surgeons to collect clinical and demographic information from participants included in the hemodynamic analysis. The collected parameters included sex (SNOMED CT: 263495000), age (SNOMED CT: 424144002), systolic blood pressure (SNOMED CT: 399304008), diastolic blood pressure (SNOMED CT: 446226005), heart rate (SNOMED CT: 399017001), smoking history (SNOMED CT: 65568007), alcohol consumption history (SNOMED CT: 219006), history of hypertension (SNOMED CT: 161501007), history of diabetes mellitus (SNOMED CT: 73211009), family history of intracranial aneurysm (SNOMED CT: 10624451000119103), portal hypertension (SNOMED CT: 34742003), liver cirrhosis (SNOMED CT: 19943007), fatty liver disease (SNOMED CT: 1231824009), and hepatitis (SNOMED CT: 128241005). In addition, the investigators recorded the presenting symptoms at the time of initial diagnosis and any history of other aneurysms.

### CTA data collection

Abdominal CT scans in this study included both conventional CT and CTA. All scans were performed using one of the following devices: a Philips Brilliance 128-slice CT scanner (Philips, Netherland), a Siemens Force dual-source CT scanner (Siemens, German), or a United Imaging uCT960+ scanner (United Imaging, China). The scanning parameters were set as follows: slice thickness of 1.0mm, slice spacing of 0.625mm, tube voltage of 100kV, tube current of 500mA, field of view of 23cm × 23cm, and matrix size of 512 × 512. Two experienced radiologists reviewed the CTA images and collected the CTA images and report information for subsequent analysis.

### Artery segmentation and 3D reconstruction

CTA images of each subject were imported into DetecModeling V02.01.16.01 software (Boea Wisdom, China) in DICOM format for vessel segmentation and 3D reconstruction [13]. First, the abdominal aorta and its major branches were automatically segmented based on the abdominal CTA sequences. Subsequently, aorta-specific geometric models were manually reconstructed, followed by smoothing and hollowing processes to generate 3D image models of the local abdominal aorta (including the CA, SMA, and bilateral renal arteries). For cases with complex anatomical structures, the research assistants collaborated with radiologists to manually refine the regions of interest using segmentation tools, ensuring the accuracy and consistency of the reconstructions. The final 3D models were exported in STL format.

Two radiologists independently reviewed the anatomical accuracy of the reconstructed models. In cases where discrepancies in aneurysm boundary identification or segmentation errors were observed, corrections were made through re-segmentation to ensure that the final models accurately reflected the vascular structures and/or the morphological characteristics of the SAAs.

### Morphological parameters collection

A total of 12 morphological parameters were selected for in-depth analysis in this study, as previous studies have demonstrated their significant association with aneurysm development and progression [14–17].

After obtaining the 3D reconstruction models, morphological parameters were extracted automatically or manually using DetecModeling V02.01.16.01 software (Boea Wisdom, China). These parameters included three vessel cross-sectional area parameters (*C_pSA_*, *C_dSA_*, *C_SMA_*), two two-dimensional size metrics (*C_SAA-N_*, *C_SAA_*), two one-dimensional size metrics (*M_L_*, *M_D_*), and three bifurcation angle parameters (*θ_SAA-SA_*, *γ_SA-pA_*, *γ_SMA-aA_*). In addition, two derived variables (*TI* and *C_pSA_/C_SMA_*) were calculated and analyzed. The specific definitions and measurement locations for each parameter are detailed in **Table 2** and **Figure 2**. Each manually measured morphological parameter was independently measured twice by standardized trained researchers, and the average value was used as the final data for analysis.

**Figure 2.**
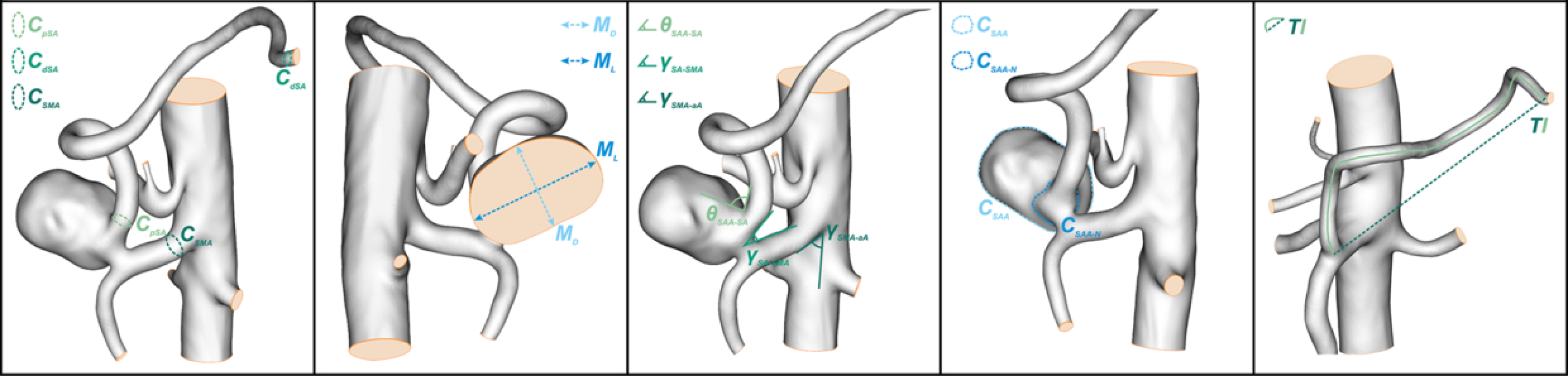
Schematic illustration of local SA morphological parameter measurements. Measurement locations for proximal and distal SA cross-sectional areas (*C_pSA_*, *C_dSA_*), SMA cross-sectional area (*C_SMA_*), aneurysm sac and neck cross-sectional areas (*C_SAA_*, *C_SAA-N_*), maximum diameter (*M_D_*) and maximum length (*M_L_*), branching angles (*θ_SAA-SA_*, *γ_SA-pA_*, *γ_SMA-aA_*), and tortuosity index (*TI*). SA, splenic artery. SAA, splenic artery aneurysm. SMA, superior mesenteric artery.

**Table 2.** Morphological parameters summary for the patients. CA, celiac axis. SA, splenic artery. SAA, splenic artery aneurysm. TI, tortuosity index.

| Table 2. Morphological parameters summary for the patients |  |
| --- | --- |
| Morphological parameters | Description |
| Vascular parameters |  |
| $C_{pSA}$ | Cross-sectional area of proximal SA |
| $C_{dSA}$ | Cross-sectional area of distal SA |
| $C_{SMA}$ | Cross-sectional area of SMA |
| 2D Shape metrics |  |
| $C_{SAA-N}$ | Maximal Cross-sectional area of SAA neck |
| $C_{SAA}$ | Maximal cross-sectional area of SAA |
| 1D Size metrics |  |
| $M_L$ | Maximal SAA length parallel to the aneurysm neck |
| $M_D$ | Maximal SAA diameter perpendicular to ML |
| Bifurcation angles |  |
| $\theta_{SAA-SA}$ | Angle between SAA and SA |
| $\gamma_{SA-pA}$ | Angle between SA and its parent artery (SMA or CA) |
| $\gamma_{SMA-aA}$ | Angle between SMA and abdominal aorta |
| Derived variables |  |
| $TI$ | The ratio of the actual vessel length to the straight-line distance |
| $C_{pSA}/C_{SMA}$ | The ratio of cross-sectional area between proximal SA and SMA |

### CFD analysis

The initial 3D models in STL format were imported into DetecFluid V01.05.14.01 software (Boea Wisdom, China) for further optimization [18]. After processing including denoising, meshing, cutting, and smoothing, final 3D models were generated for CFD analysis. The mesh size was set to 0.2mm, producing approximately 400,000 finite volume elements, which ensured high mesh quality with favorable triangle aspect ratios and uniform element sizes, thereby facilitating computational stability and convergence.

The Navier-Stokes equations were discretized using a second-order upwind scheme and a first-order implicit Euler scheme. Blood was modeled as an incompressible Newtonian fluid with a density of 1,060 kg/m^3^ and a dynamic viscosity of 0.004Pa·s, and the vessel wall was assumed to be rigid with a no-slip boundary condition [19–23]. The outlets of the branch vessels were coupled with a three-element Windkessel model to simulate the impedance characteristics of the downstream vasculature [24].

To determine the Windkessel model parameters (characteristic impedance Rc, peripheral resistance Rp, and compliance C) for each outlet, we first estimated the initial flow distribution based on the cross-sectional areas of each branch vessel measured from CTA images, combined with physiologically reported flow partitioning ratios [25,26]. An iterative optimization algorithm was then applied to adjust the Rp values at each outlet, with patient-specific cardiac output, systolic blood pressure, and diastolic blood pressure as constraints, until the calculated mean flow at each outlet matched the initial flow distribution targets and the overall pressure waveform aligned with clinical measurements. The final calibrated Windkessel parameters ensured that the systolic blood pressure, diastolic blood pressure, and flow distribution at each outlet obtained from the 3D simulations were consistent with clinical data [27,28]. The inlet velocity waveform was adapted from a typical ascending aortic flow waveform to match patient-specific hemodynamic parameters, including cardiac output, heart rate, and the systolic-to-diastolic duration ratio [29].

The computational time step was fixed at 10ms. At each time step, convergence was considered achieved when the normalized residuals fell below 10^-4^ [30]. Transient CFD simulations were run for 10 cardiac cycles to ensure a periodic steady state, and subsequent analyses were based on the results of the final cycle.

### Hemodynamic parameters collection

A total of 14 hemodynamic parameters across two categories were extracted from the simulated flow fields, which have been recognized as key determinants in aneurysm initiation and progression [28–30].

The first category comprised seven parameters related to flow velocity and flow rate: mean flow velocity at the proximal SMA (m/s), mean flow velocity at the proximal CA (m/s), mean flow velocity at the proximal SA (m/s), mean flow velocity at the distal SA (m/s), flow rate at the proximal SA (mL/s), flow rate at the distal SA (mL/s), and flow rate at the parent artery of the SA (mL/s).

The second category included six parameters related to mean pressure and WSS: mean pressure at the SAA (mmHg), mean pressure at the SA (mmHg), mean pressure at the proximal SA (mmHg), mean pressure at the distal SA (mmHg), time-averaged wall shear stress (TAWSS) at the proximal SA (Pa), and TAWSS at the distal SA (Pa).

In addition, one dimensionless parameter, the OSI, was incorporated.

Each hemodynamic parameter was independently measured twice by standardized trained researchers, and the average value of the two measurements was used as the final data for analysis.

### Machine learning model development

This study employed a two-stage prediction strategy to jointly model the prediction of SAA occurrence and localization, simulating the real-world clinical decision-making process: first determining whether a patient has an aneurysm, and if so, further identifying its specific location.

#### Stage 1: Occurrence Prediction

Given the relatively limited sample size of 195 patients, we implemented a stringent feature selection strategy to mitigate the risk of model overfitting and ensure generalizability. Although a total of 12 morphological parameters were initially measured, we subsequently refined the feature set based on multicollinearity diagnostics (variance inflation factor <5) and clinical interpretability. In addition, hemodynamic parameters were deliberately excluded from the initial model input to prevent causal confounding, as these parameters are themselves downstream manifestations of morphological geometry and are collinear with the anatomical features. Consequently, five morphological parameters (*C_dSA_*, *γ_SA-pA_*, *γ_SMA-aA_*, *C_pSA_/C_SMA_*, and *TI*) were ultimately selected as the core predictors. These parameters were chosen because they demonstrated the strongest independence in the correlation matrix and the highest discriminatory power in univariate analyses. No missing values were present in the morphological parameters used for model training. No outliers were identified or removed, all samples were retained for model training.

Supervised learning was employed for both the occurrence prediction and location prediction tasks. Based on five morphological parameters, nine ML algorithms were incorporated to construct binary classification models for SAA prediction (occurrence vs. non-occurrence), including logistic regression, random forest, linear kernel support vector machine (SVM), K-nearest neighbors (KNN), decision tree, gradient boosting decision tree (GBDT), Naïve Bayes, linear discriminant analysis (LDA), and extreme gradient boosting (XGBoost). These models were selected due to their favorable generalization performance and interpretability in small-sample medical prediction tasks. Logistic regression, being the most commonly used modeling technique in traditional risk prediction, was employed as the baseline comparison model. All features were standardized using Z-score normalization to eliminate the effects of different scales.

Given the relatively small sample size (n=195), leave-one-out cross-validation (LOOCV) was adopted for model evaluation. This approach uses n-1 samples for training and the remaining single sample for validation at each iteration, representing one of the most reliable evaluation strategies for small-sample datasets. Principal component analysis (PCA) was performed to assess the performance differences between models built on original and reduced features, with the number of principal components determined based on a cumulative variance explanation rate of ≥90%. Preliminary analysis revealed that the dataset comprised 94 cases (48.2%) with aneurysms and 101 cases (51.8%) without aneurysms, indicating a relatively balanced class distribution; therefore, no oversampling techniques were applied.

Model performance was assessed using the following metrics: area under the receiver operating characteristic curve (AUROC) with 95% confidence intervals (CIs), accuracy, sensitivity, specificity, positive predictive value (PPV), and negative predictive value (NPV). The 95%CIs of the AUROC were calculated using the Bootstrap method with 2,000 resampling iterations. Platt scaling was applied to calibrate the predicted probabilities of the optimal model, and calibration performance was evaluated using the Brier score. Calibration curves were plotted to assess the agreement between predicted probabilities and actual observed outcomes, and decision curve analysis (DCA) was performed to evaluate the clinical net benefit of the models across different threshold probabilities. Among the models compared, the KNN model achieved the best performance, and Platt scaling was further applied to calibrate its predicted probabilities to enhance clinical applicability. Feature importance was quantified using SHapley Additive exPlanations (SHAP) to improve model interpretability.

#### Stage 2: Location Prediction

For samples predicted as ‘aneurysm occurrence’ in the first stage, a location prediction model (multiclass classification: proximal SA vs. distal SA) was further constructed. Random forest was employed as the base learner, with accuracy and macro-averaged F1-score as the primary evaluation metrics. This two-stage strategy simulates the actual clinical decision-making pathway by first determining the presence of the disease (qualitative) and then identifying its location (localization), thereby offering favorable clinical interpretability.

All statistical analyses and ML modeling were performed using Python 3.14, with core libraries including scikit-learn, SHAP, and Matplotlib. A two-sided *P*<0.05 was considered statistically significant.

### Web application development

In this study, the optimal ML models (the occurrence prediction model and the location prediction model) were encapsulated into an interactive web application to facilitate their clinical translation and application. The application was developed using the Flask framework (version 3.0.0), with the backend API service implemented in Python and the frontend interface built with HTML5, CSS3, and native JavaScript, supporting bilingual switching between Chinese and English.

The prediction workflow of the web application comprises two stages: (1) users input five morphological parameters (*C_pSA_/C_SMA_*, *C_dSA_*, *γ_SA-pA_*, *γ_SMA-aA_*, and *TI*) on the frontend interface; (2) the frontend sends the parameters as a JSON payload to the backend API via an HTTP POST request; (3) upon receiving the parameters, the backend calls a preloaded standard scaler to normalize the input data and feeds them into the trained KNN model for occurrence probability prediction; (4) if the prediction result is negative (no aneurysm), a low-risk result with follow-up recommendations is directly returned; (5) if the prediction result is positive (aneurysm present), the backend automatically invokes the preloaded random forest location prediction model to further predict the aneurysm location (proximal SA vs. distal SA), and returns both the location prediction and the occurrence probability to the frontend; and (6) the frontend dynamically renders the prediction results, including occurrence probability, confidence level, risk stratification, predicted location, and corresponding clinical recommendations.

The model was deployed using a client-server architecture. The backend service was run using the Flask built-in development server. All model files, including the trained KNN occurrence prediction model, the random forest location prediction model, the standard scaler, and the feature name list, were stored in Pickle format (.pkl) and preloaded into memory upon service startup to ensure real-time prediction responses. The frontend interface was designed with a responsive layout, compatible with both desktop and mobile browsers, facilitating use by clinicians across various devices. The application has been deployed on an Alibaba Cloud server and is remotely accessible at http://zs-vascular.asia/ (also available at http://8.153.147.47:5000/). A schematic diagram of the application architecture is presented in **Figure 1C**.

### Independent data access and responsibility

Authors had full access to all the data in this study and takes full responsibility for the integrity of the data and the accuracy of the data analysis. The source code and model files of this web application have been open-sourced on GitHub (https://github.com/RanXu1995/zs_abeSA-model) to facilitate reproducibility and further optimization by the research community.

### Statistical analysis

Statistical analyses were performed using R software (version 4.5.2), and data visualization was conducted using the ggplot2 package. Normality of continuous variables was assessed using the Shapiro-Wilk test. Normally distributed continuous variables were presented as mean ± standard deviation, while non-normally distributed variables were presented as median (interquartile range). Homogeneity of variances was tested using Levene’s test. For variables that were normally distributed with equal variances, the independent samples t-test was used; for variables that were not normally distributed, the Mann-Whitney U test was used; and for variables that were normally distributed but with unequal variances, Welch’s t-test was used. For comparisons of categorical data with small sample sizes (n<40), Fisher’s exact test was applied. Correlation analysis between two non-normally distributed continuous variables was performed using Spearman’s rank correlation test. All hypothesis tests were two-sided, and a *P*<0.05 was considered statistically significant.

## Results

### Clinical baseline characteristics

Aberrant SA is defined as a congenital anatomical variation in which the SA arises from the SMA (**Figure 3A**). To investigate the pathogenesis of this condition, we enrolled 195 patients with aberrant SA anatomy from 16 centers between January 2008 and June 2026 (**Figure 3B**, **Table 3**). Among them, 94 patients developed SAAs in the setting of this aberrant anatomy, while 101 did not. The mean age of the patients was 59.0±14.7 years (range: 27-85 years), and female patients accounted for 48.7% (n=95). In addition, a cohort of patients with SAAs without anatomical anomalies (n=100) and a cohort of individuals without a history of abdominal aortic disease (n=100) were enrolled as control groups. Notably, only 18.5% (n=36) of patients with aberrant SA anatomy presented with obvious clinical symptoms (33.0% in the aberrant SA with aneurysm group vs. 5.0% in the aberrant SA without aneurysm group). Among these, 28 patients complained of abdominal pain, and 6 complained of abdominal distension. In addition, two patients with aberrant SAAs were diagnosed after presenting with acid reflux, heartburn, and fatigue.

**Figure 3.**
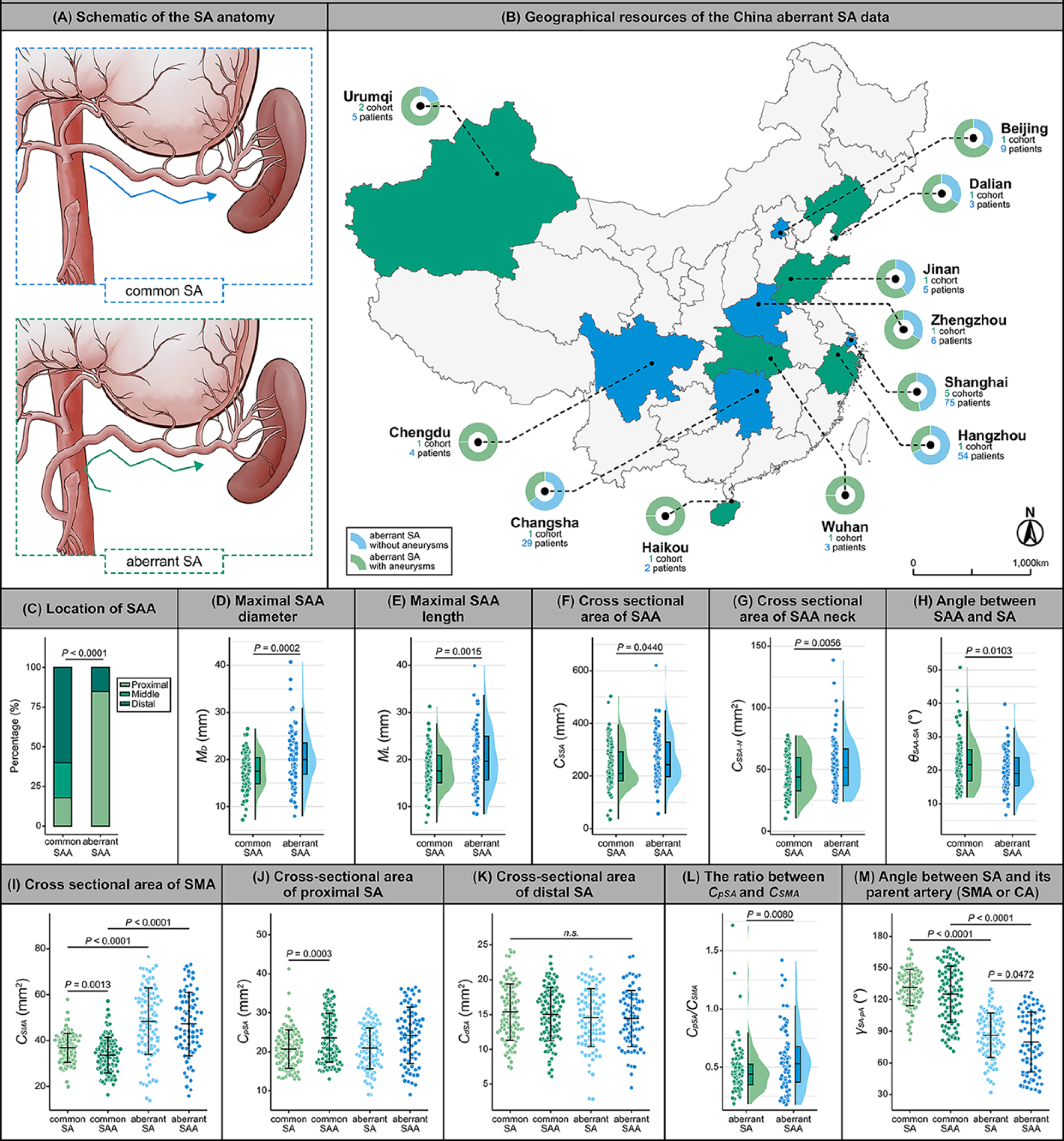
Baseline characteristics and morphological analysis of patients with aberrant SA anatomy. **(A)** Schematic illustration of aberrant SA anatomy. (B) Geographic distribution of the study cohort (n=195). **(C)** Comparison of anatomical location distribution between aberrant SAAs and common SAAs. **(D-G)** Comparison of morphological parameters between aberrant SAAs and common SAAs: maximum diameter **(D)**, maximum length **(E)**, maximum aneurysm sac cross-sectional area **(F)**, and maximum aneurysm neck cross-sectional area **(G)**. **(H)** Comparison of the angle between the aneurysm sac and the main axis of the SA. **(I-M)** Comparison of local vascular morphological parameters among the four groups: SMA cross-sectional area **(I)**, proximal and distal SA cross-sectional areas **(J,K)**, SA-to-SMA cross-sectional area ratio **(L)**, and SA-to-parent vessel branching angle **(M)**. SA, splenic artery. SAA, splenic artery aneurysm. SMA, superior mesenteric artery.

**Table 3.**
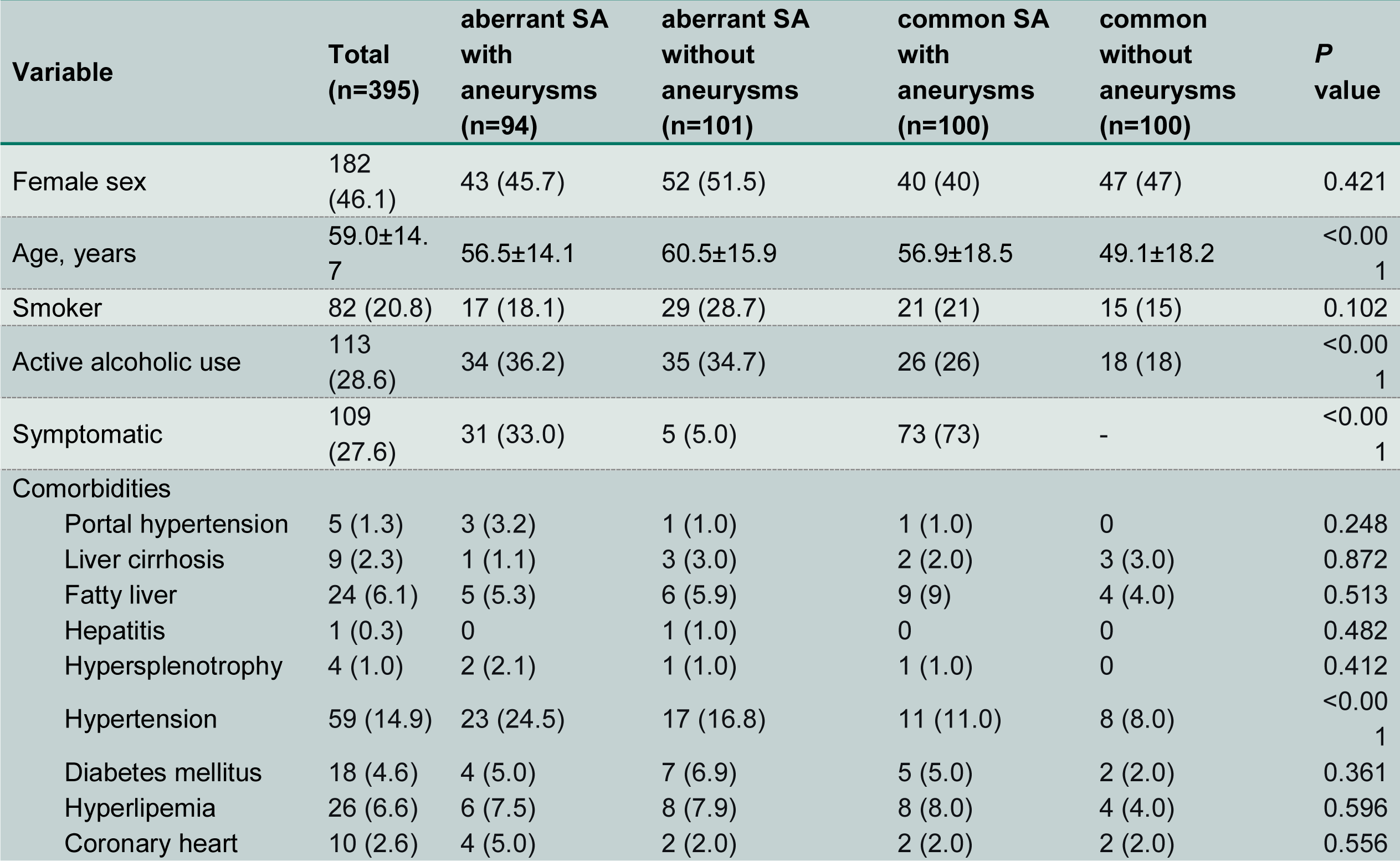
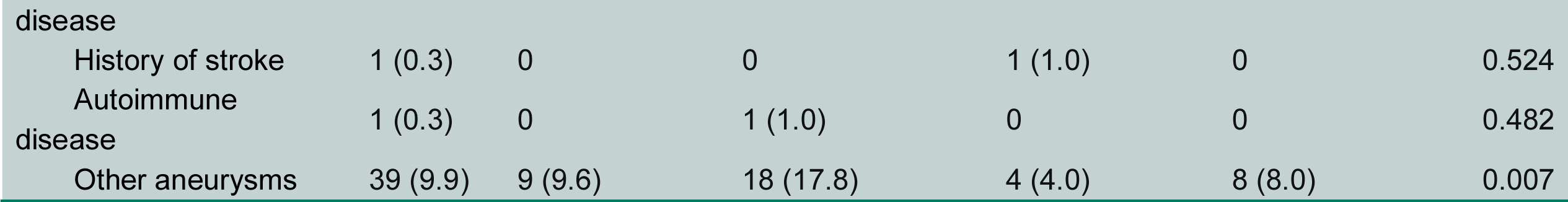
Demographics and comorbidities of the patients. Categorical variables are presented as number (%). Continuous variables are presented as mean ± standard deviation. SA, splenic artery. SAA, splenic artery aneurysm.

Comorbidities in patients with aberrant SA anatomy were statistically analyzed. Twenty-five patients (12.8%) had concomitant liver diseases, including fatty liver disease in 11 cases (5.6%), portal hypertension combined with liver cirrhosis in 3 cases (1.5%), fatty liver disease combined with liver cirrhosis in 10 cases (5.1%), and hepatitis in 1 case (0.5%). Furthermore, 40 patients (20.5%) had hypertension, 11 (5.6%) had diabetes mellitus, 14 (7.2%) had hyperlipidemia, and 6 (3.1%) had coronary artery disease. Twenty-seven patients (13.8%) had aneurysms at other locations, including 17 patients with abdominal aortic aneurysms (all of whom had undergone resection and prosthetic graft replacement) and 10 patients with other VAAs (including SMA aneurysms and hepatic artery aneurysms). It should be noted that this proportion may be subject to selection bias, as all enrolled patients in this study underwent abdominal CTA based on clinical indications.

### Morphological characteristics of aberrant SAA

We analyzed 101 SAAs in 94 patients with aberrant SA anatomy and found that the majority of patients (n=89, 94.7%) had a solitary SAA, while 5 patients had multiple SAAs in the setting of an aberrant SA. Regarding anatomical location, the vast majority of aberrant SAAs were located in the proximal SA (n=80, 85.1%), whereas only 14.9% (n=14, including 5 cases of multiple aneurysms) occurred in the distal/splenic hilar region. No aneurysm located in the middle segment of the SA was observed in this cohort (**Figure 3C**). In contrast, the common primary SAA cohort (n=100) exhibited a distinctly different distribution pattern, with aneurysms more frequently located in the distal splenic hilum (60.2%), while the proportions at the proximal and middle segments were relatively lower (15.5% and 16.3%, respectively). This distribution pattern is consistent with previous literature reports [27,31].

In addition to the location distribution analysis, we performed vessel segmentation and 3D reconstruction of CTA data from all four groups of patients. Based on the reconstructed 3D models, we were able to measure the local morphological parameters of the SA and SAAs more accurately and conveniently. Our results demonstrated that, compared with common SAAs, aberrant SAAs had a larger maximum diameter (20.4mm vs. 17.5mm, *P<*0.001, **Figure 3D**) and a greater maximum length (20.5mm vs. 17.9mm, *P*=0.002, **Figure 3E**). In addition, the maximum cross-sectional areas of both the aneurysm sac and the aneurysm neck were significantly larger in aberrant SAAs than in common SAAs (sac: 263.8mm^2^ vs. 236.7mm^2^, *P*=0.044; neck: 54.3mm^2^ vs. 46.2mm^2^, *P*=0.006, **Figure 3F,G**). Further analysis of the angle between the aneurysm sac and the main axis of the SA revealed that the angle was smaller in aberrant SAAs than in common SAAs (22.3° vs. 19.7°, *P*=0.010, **Figure 3H**). A smaller angle between the aneurysm sac and the parent vessel may result in stronger hemodynamic impact on the aneurysm sac surface, thereby increasing wall pressure, which may explain why aberrant SAAs tend to be larger than common SAAs.

Subsequently, we performed a comparative analysis of local vascular morphological parameters across all four groups. As shown in **Figure 3I**, compared with normal anatomy, when the SA arose from the SMA, the mean cross-sectional area of the SMA (from the aortic bifurcation to the SA bifurcation) was significantly increased (non-aneurysm group: *P*<0.001; aneurysm group: *P*<0.001). However, no significant changes were observed in the mean cross-sectional area of the SA, either proximally or distally, compared with normal anatomy (**Figure 3J,K**). Since the aberrant SA arises from the SMA, we further calculated the ratio of the SA to SMA cross-sectional areas in the non-aneurysmal and aneurysmal aberrant SA groups. Interestingly, this ratio was significantly higher in the aberrant SAA group than in the non-aneurysm group (0.57 vs. 0.47, *P*=0.008, **Figure 3L**). A higher ratio suggests that the SA may receive a greater blood flow volume, thereby increasing local vascular wall pressure.

In addition, we analyzed the angular relationships between the various vessels. We found that the angle between the SA and its parent vessel (the CA or the SMA) differed significantly among the groups. As shown in **Figure 3M**, when the SA arose from the SMA, the angle between the SA and its parent vessel was significantly smaller than when it arose from the CA (non-aneurysm group: *P*<0.001; aneurysm group: *P*<0.001). This phenomenon was even more pronounced in the aberrant SAA group (86.3° vs. 78.8°, *P*=0.047). These differences in morphological parameters may promote the development and progression of SAAs in the setting of aberrant SA anatomy by influencing local hemodynamics.

### Follow-up outcome of non-aneurysmal aberrant SAA

We followed up 101 patients with aberrant SA anatomy who did not develop SAAs to assess their potential risk of future aneurysm progression. Effective follow-up data were ultimately obtained from 14 patients across three centers, with a mean follow-up duration of 36.8±22.1 months (range: 7-76 months) and a mean of 2.9±0.8 follow-up visits. Since no middle-segment aberrant SAAs were observed in this cohort, the observation region was focused on the proximal SA and the distal/splenic hilar region. **Figure 4A** shows CTA images of two representative patients who were followed up for 62 and 76 months, respectively. The results showed no evident expansion over time in either the diameter of the proximal SA or the vessel diameter at the distal/splenic hilar region. Linear mixed-effects model analysis of imaging data from all 14 followed patients further demonstrated that both the diameters and cross-sectional areas of the proximal and distal SA remained stable during the follow-up period, with no significant increasing trend (diameter-proximal: β=0.0033mm/month, 95%CI: -0.0006-0.0072, *P*=0.094; distal/splenic hilum: β=0.0004mm/month, 95%CI: -0.0003-0.0017, *P*=0.703. Cross-sectional area-proximal: β=0.0138mm^2^/month, 95%CI: -0.0836-0.1112, *P*=0.780; distal/splenic hilum: β=0.0072mm^2^/month, 95%CI: -0.0452-0.0596, *P*=0.786) (**Figure 4B-E**).

**Figure 4.**
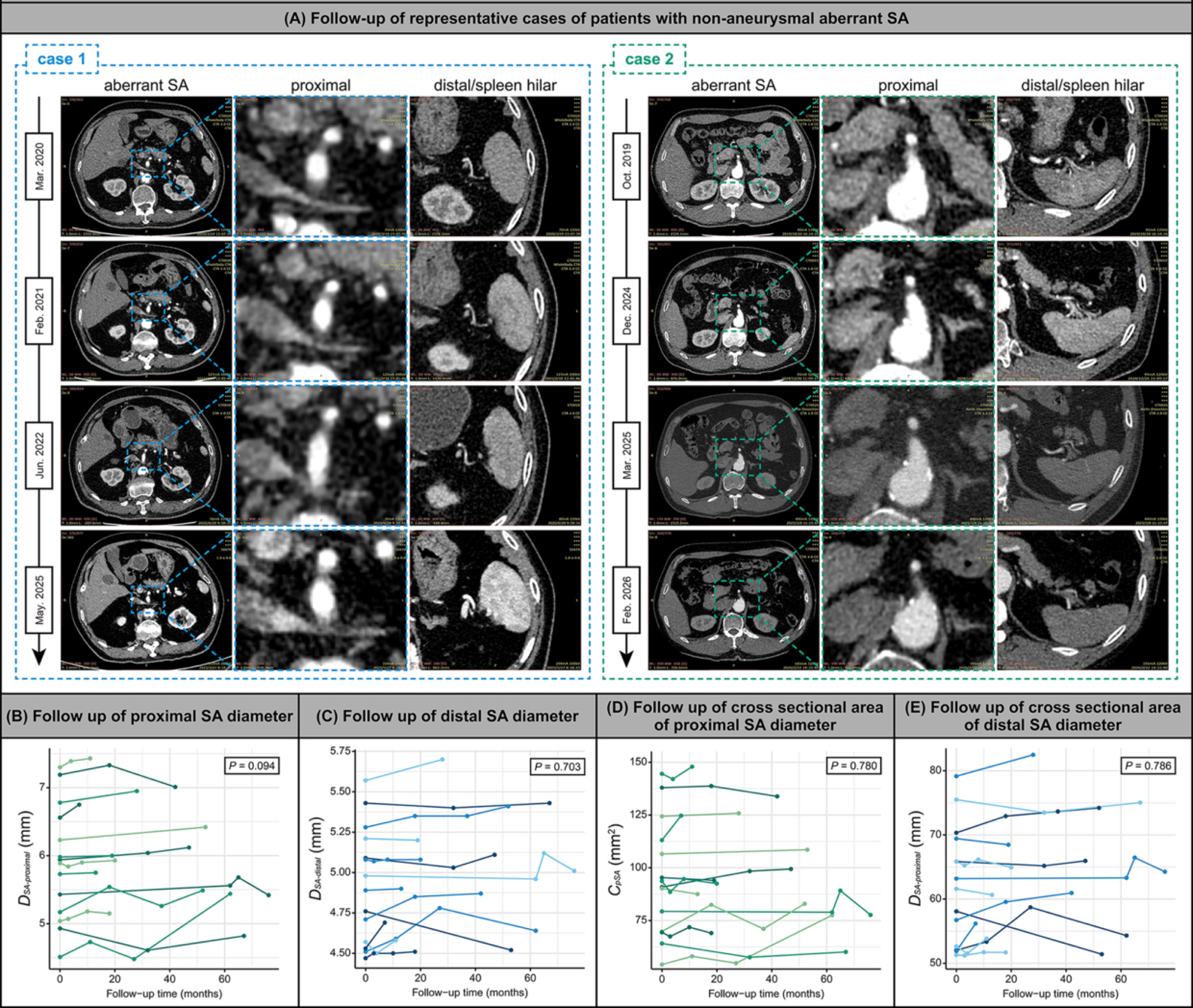
Follow-up analysis of non-aneurysmal patients with aberrant SA anatomy. **(A)** Serial CTA follow-up images of two representative patients (follow-up at 62 and 76 months), demonstrating stable proximal and distal SA diameters over time. **(B-E)** Linear mixed-effects model analysis of proximal and distal SA diameters **(B,C)** and cross-sectional areas **(D,E)** over time, showing no significant enlargement (all *P*>0.05). Each measurement was performed three times, and measured data are presented as the mean±SD. SA, splenic artery. SAA, splenic artery aneurysm.

These findings suggest that the development of aberrant SAAs does not arise from a chronic degenerative pathological process, but rather from local hemodynamic alterations induced by specific unfavorable morphological anatomical structures, with the process likely being acute or subacute in nature. In other words, by identifying adverse morphological features and their associated hemodynamic changes, we can screen for high-risk individuals in the context of aberrant SA anatomy and predict their future risk of SAA formation.

### Blood flow patterns of aberrant SAA

Based on the reconstructed 3D models (**Figure 5A**) and patient clinical data (e.g., systolic/diastolic blood pressure, stroke volume, etc.), we performed CFD analysis of the local abdominal aorta region for each patient (**Table 4**). As shown in the time-averaged velocity streamline maps (**Figure 5B**) and velocity vector maps (**Figure 5C**), compared with normal vascular regions, blood flow within the SAA segment exhibited pronounced disturbances, including low-velocity vortices and helical turbulence. Low-velocity regions were mainly located in the distal SA of common SAAs and the proximal lateral side of the SA in aberrant SAAs, whereas high-velocity flow tended to be distributed on the medial side. Further quantitative analysis of cross-sectional flow velocities in major branch vessels (proximal SMA, proximal CA, and proximal and distal SA) revealed that the proximal flow velocity of the SA in the aberrant anatomy was significantly decreased, particularly in the aberrant SAA group (aberrant SAA vs. common SAA: 0.08m/s vs. 0.14m/s, *P*<0.001; aberrant SAA vs. non-aneurysmal aberrant SA: 0.08m/s vs. 0.12m/s, *P*<0.001, **Figure 5D,E**). In contrast, the distal flow velocity of the SA in patients with aberrant SAAs was significantly increased compared with the common SA group, especially in the aberrant SAA group (aberrant SAA vs. common SAA: 0.41m/s vs. 0.34m/s, *P*<0.001, **Figure 5F**). Quantitative analysis of flow velocity within the parent vessel supplying the SA (i.e., the CA in normal anatomy and the SMA in patients with aberrant SA) revealed no statistically significant differences (**Figure 5G**).

**Figure 5.**
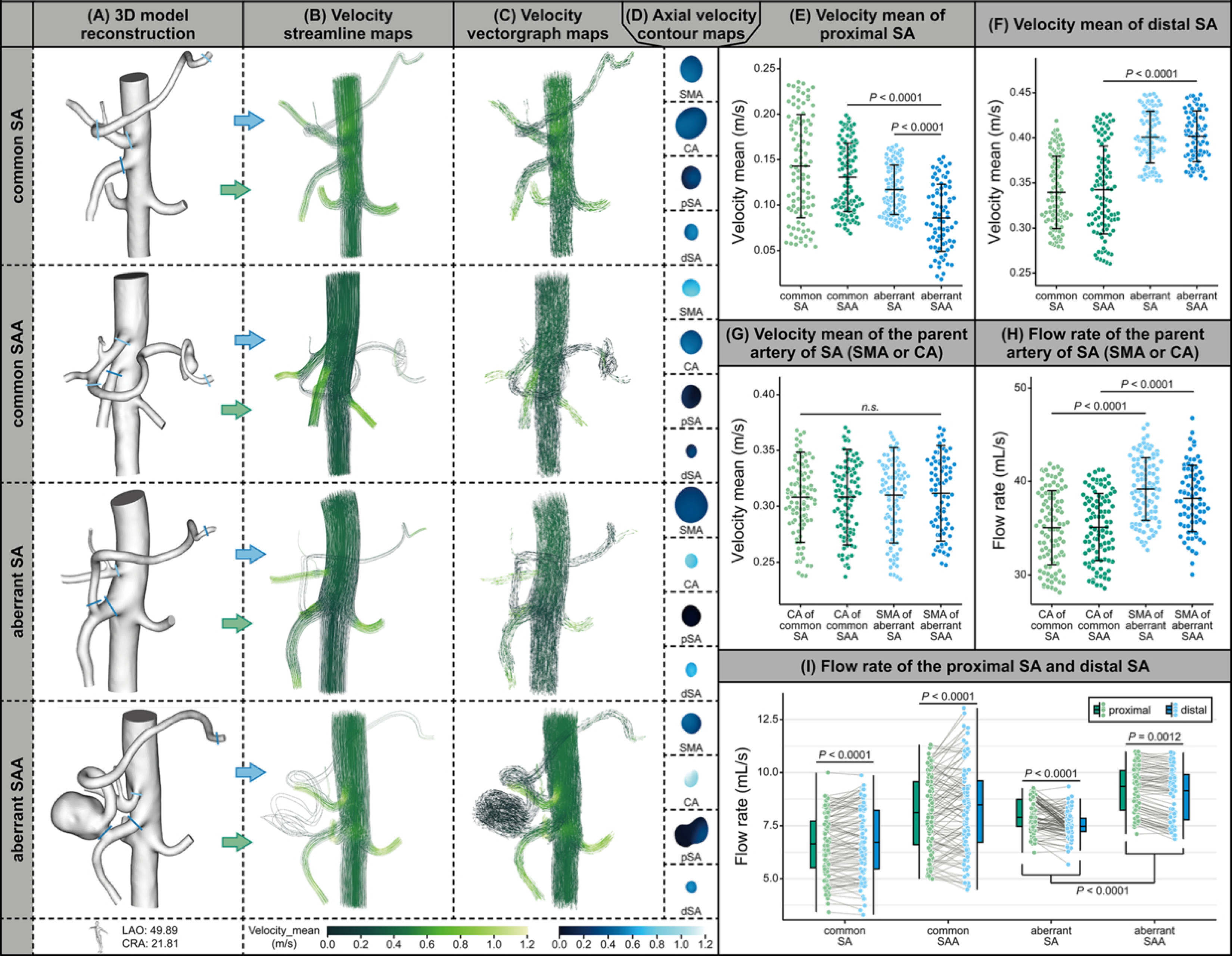
Hemodynamic analysis of flow velocity and flow rate. (A-D) 3D reconstruction models **(A)**, time-averaged velocity streamline maps **(B)**, velocity vector maps **(C)**, and schematic of cross-sectional velocity quantification in major branch vessels **(D)** of the four groups. **(E-G)** Comparison of proximal SA velocity **(E)**, distal SA velocity **(F)**, and parent vessel velocity **(G)** among the groups. **(H)** Comparison of flow rate between the parent artery of SA (SMA or CA). **(I)** Comparison of proximal and distal SA flow rates among the groups. Celiac axis, CA. SA, splenic artery. SAA, splenic artery aneurysm. SMA, superior mesenteric artery. Each measurement was performed three times, and measured data are presented as the mean±SD.

**Table 4.**
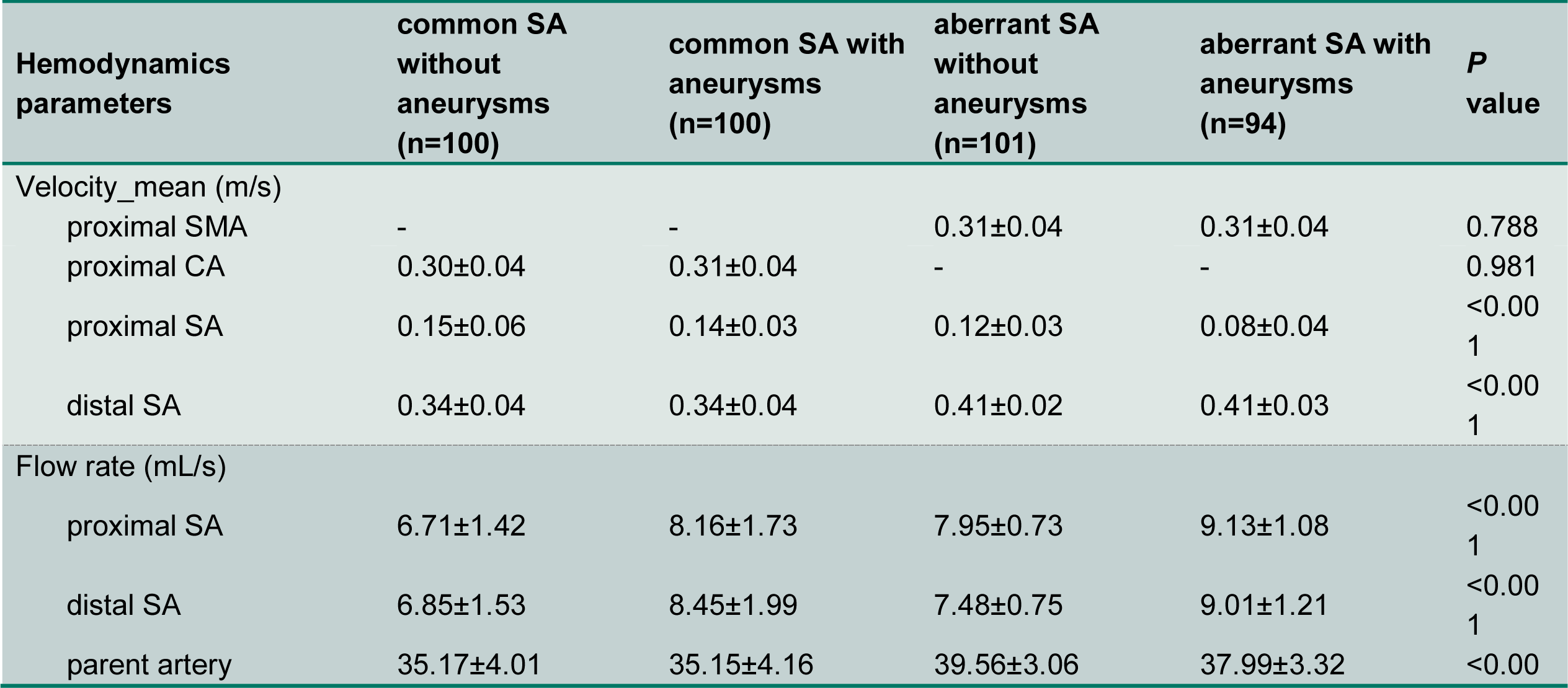
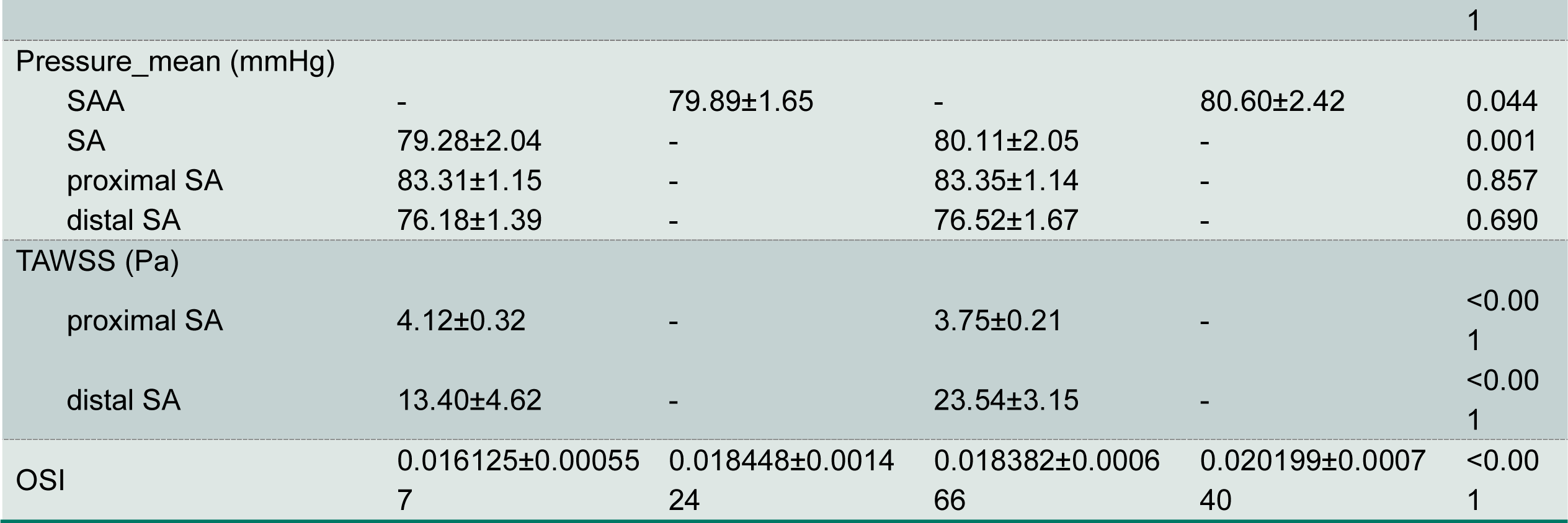
Hemodynamic parameters summary for the patients. Continuous variables are presented as mean ± standard deviation. CA, celiac axis. SA, splenic artery. SAA, splenic artery aneurysm. TAWSS, time-averaged wall shear stress. OSI, oscillatory shear index.

Cross-sectional flow rate within the lumen was calculated by combining cross-sectional flow velocity and cross-sectional area. Flow rate directly affects vascular wall pressure. As shown in **Figure 5H**, in the aberrant SA anatomy, the flow rate of the SMA was significantly higher than that of the CA in normal anatomy (non-aneurysm group: *P*<0.001; aneurysm group: *P*<0.001), which may result in the SA receiving a greater blood flow volume and, consequently, increased internal pressure. We also observed that in normal anatomy, the distal SA flow rate was higher than the proximal flow rate (non-aneurysm group: *P*<0.001; aneurysm group: *P*<0.001), whereas in the aberrant SA anatomy, the proximal SA flow rate was higher than the distal flow rate (non-aneurysm group: *P*<0.001; aneurysm group: *P*=0.001). Moreover, the SA flow rate at all locations in patients with aberrant SAAs was significantly higher than that in patients without aneurysms (**Figure 5I**). The higher flow rate in the aberrant SAA group (especially at the proximal segment) may be related to its lower flow velocity and larger cross-sectional area. This finding well explains our previous observation that aberrant SAAs are more likely to occur in the proximal segment.

### General hemodynamics characteristics of aberrant SAA

Based on the 3D reconstruction models, we also analyzed the local pressure isopleth distribution of the abdominal aorta (**Table 4**). As shown in **Figure 6A**, focal high pressure was observed in the local aneurysm region of both common SAAs and aberrant SAAs. Through segmental extraction and quantitative analysis of the entire SA, we found that the local mean pressure of aberrant SAAs was slightly higher than that of common SAAs (80.60mmHg vs. 79.89mmHg, *P*=0.044, **Figure 6E**), and the mean pressure of the entire SA harboring the aneurysm was also significantly higher in aberrant SAAs than in common SAAs (80.11mmHg vs. 79.28mmHg, *P*=0.001, **Figure 6F**). This may further explain why aberrant SAAs tend to have larger dimensions. However, quantitative analysis of mean pressure in the proximal and distal segments of the SA between normal anatomy and non-aneurysmal aberrant SAs revealed no statistically significant differences (**Figure 6G,H**).

**Figure 6.**
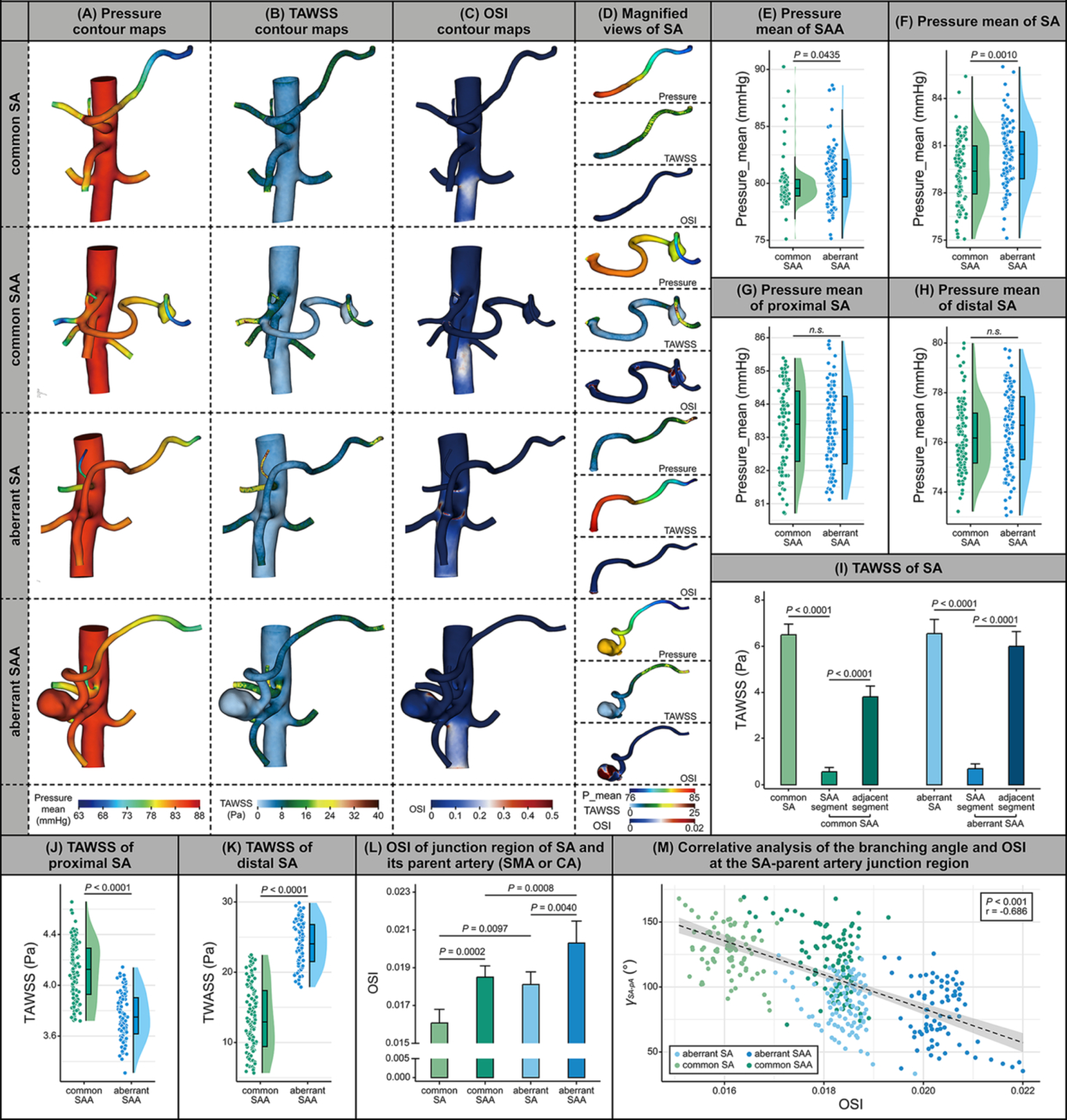
Hemodynamic analysis of pressure, WSS, and OSI. (A-C) Local pressure isopleth maps **(A)**, TAWSS contour maps **(B)**, OSI contour maps **(C)** of the four groups. **(D)** Schematic illustration of pressure quantification in the localized SA segment. **(E-H)** Comparison of local SAA mean pressure **(E)**, whole SA mean pressure **(F)**, proximal SA pressure **(G)**, and distal SA pressure **(H)** among common SAAs and aberrant SAAs. **(I)** Comparison of TAWSS among non-aneurysmal regions, aneurysm regions, and adjacent regions. **(J,K)** Comparison of proximal SA **(J)** and distal SA **(K)** TAWSS between common SAAs and aberrant SAAs. **(L)** Comparison of OSI at the SA-parent vessel junction among groups. **(M)** Correlation analysis between OSI at the SA-parent vessel junction and the SA-parent vessel branching angle among groups. SA, splenic artery. SAA, splenic artery aneurysm. SMA, superior mesenteric artery. TAWSS, time-averaged wall shear stress. OSI, oscillatory shear index. Each measurement was performed three times, and measured data are presented as the mean±SD.

In addition, we calculated the TAWSS and OSI in the local abdominal aortic region. The overall and local TAWSS contour maps are shown in **Figure 6B** and **Figure 6D**, respectively. Compared with either the regions adjacent to the aneurysm or the non-aneurysmal SA, the aneurysm region of the SA exhibited lower TAWSS (common SAA: *P*<0.001; aberrant SAA: *P*<0.001, **Figure 6I**). We further quantitatively calculated the proximal and distal TAWSS of the SA in normal anatomy and in the aberrant SA. We found that in patients with an aberrant SA arising from the SMA, the proximal TAWSS was significantly lower than that in the proximal SA arising from the CA in normal anatomy (4.12 vs. 3.75, *P*<0.001, **Figure 6J**). In contrast, the distal TAWSS was lower in the common SA arising from the CA (13.4 vs. 23.5, *P*<0.001, **Figure 6K**). These findings suggest that the anatomical variation in which the SA arises from the SMA may subject the proximal wall of the SA to a chronically low-shear-stress environment, whereas the SA arising from the CA exhibits lower distal TAWSS. This phenomenon well explains the distribution pattern of aberrant SAAs predominantly occurring in the proximal segment, while common SAAs tend to occur in the distal segment.

OSI analysis revealed that, in addition to high OSI distribution within the dilated aneurysm sac, the OSI value at the junction of the SA and its parent vessel (the CA or the SMA) was also significantly elevated (**Figure 6C,D**). Precise quantitative analysis demonstrated that patients with concomitant aneurysms had higher OSI values at the SA-parent vessel junction than those without aneurysms (common SA: *P<*0.001; aberrant SA: *P=*0.004), and the aberrant SA anatomy further exacerbated the elevation of OSI at this junction (**Figure 6L**). Integrating the aforementioned morphological finding regarding the anatomical angle between the SA and its parent vessel, we further performed a correlation analysis, which showed that the OSI value was inversely proportional to the SA-parent vessel angle, i.e., a smaller angle was associated with a higher OSI value (*P*<0.001, r=-0.686, **Figure 6M**). This indicates that in the aberrant SA anatomy, the region at the bifurcation of the SA and the SMA, as well as the lateral wall of the proximal SA, exhibits slow blood flow and a propensity for vortex formation due to the sharp angle, accompanied by the features of low TAWSS and high OSI, which is precisely the predilection site for aberrant SAAs.

### Comparative hemodynamic analysis between aberrant SA and aberrant SAA

Multiple studies have demonstrated that restoring the pre-aneurysmal morphology is of great value for elucidating the pathogenesis and disease evolution of aneurysms [32,33]. To further investigate the potential mechanisms underlying why some patients develop SAAs while others do not in the context of aberrant SA anatomy, we performed morphological restoration of the pre-aneurysmal state in patients with aberrant SAAs (**Figure 7A**) and compared their morphological and hemodynamic parameters with those of patients with aberrant SA without aneurysms.

**Figure 7.**
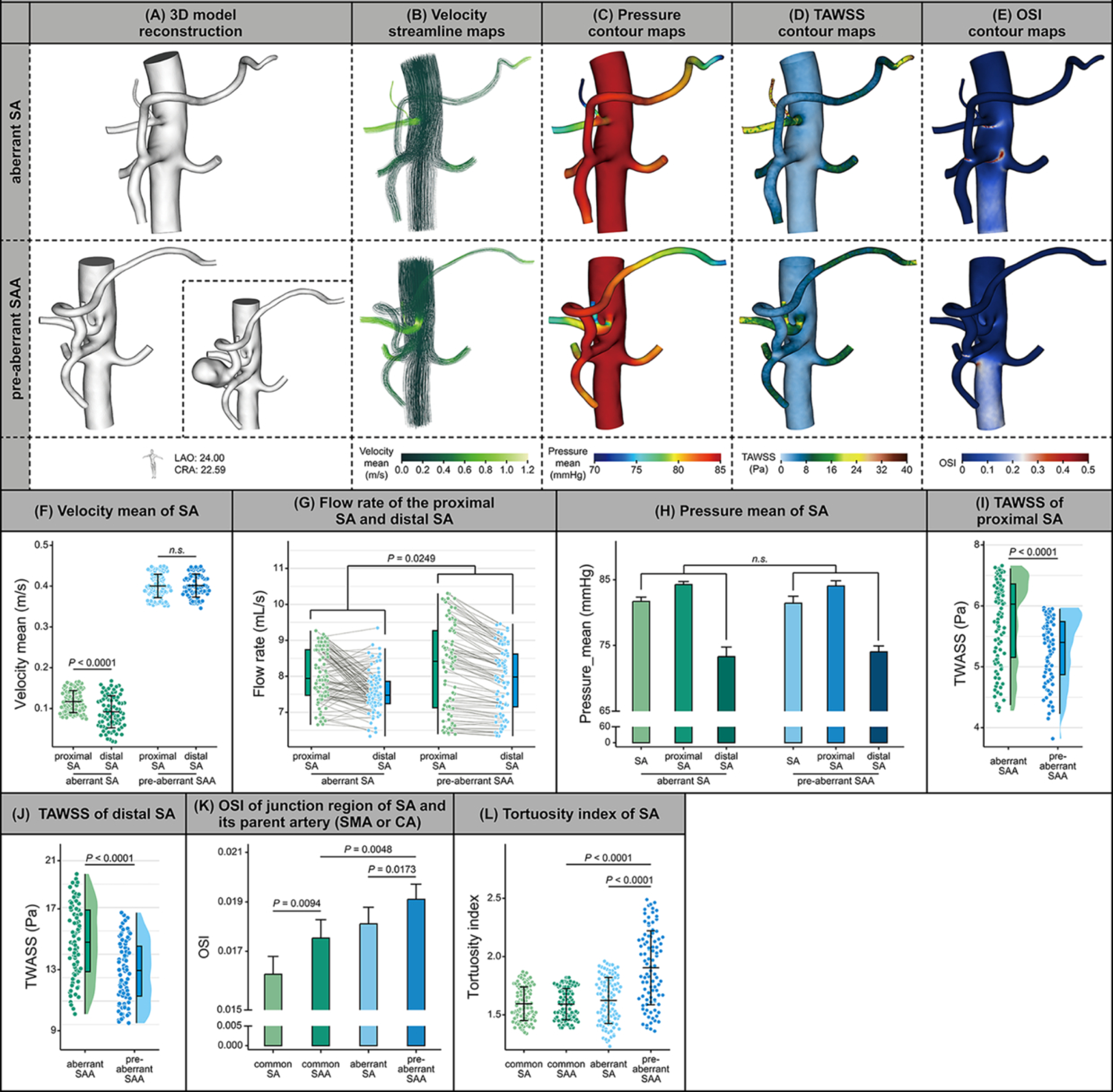
Pre-aneurysmal morphological restoration and hemodynamic analysis of aberrant SAAs. (A-E) Pre-aneurysmal morphological restoration in patients with aberrant SAAs: 3D reconstructed model **(A)**, time-averaged velocity streamline maps **(B)**, local pressure isopleth maps **(C)**, TAWSS contour maps **(D)**, and OSI contour maps **(E)**. **(F,G)** Comparison of proximal and distal SA velocity **(F)** and flow rate **(G)** between the restored pre-aneurysmal aberrant SAA group and the non-aneurysmal aberrant SA group. **(H)** Comparison of overall, proximal, and distal mean SA pressure between the restored pre-aneurysmal aberrant SAA group and the non-aneurysmal aberrant SA group. **(I,J)** Comparison of proximal **(I)** and distal **(J)** TAWSS between the restored pre-aneurysmal aberrant SAA group and the non-aneurysmal aberrant SA group. **(K)** Comparison of OSI at the SA-parent vessel junction among the normal SA group, common SAA group, restored pre-aneurysmal aberrant SAA group, and non-aneurysmal aberrant SA group. **(L)** Comparison of tortuosity index among the normal SA group, common SAA group, restored pre-aneurysmal aberrant SAA group, and non-aneurysmal aberrant SA group. SA, splenic artery. SAA, splenic artery aneurysm. SMA, superior mesenteric artery. TAWSS, time-averaged wall shear stress. OSI, oscillatory shear index. Each measurement was performed three times, and measured data are presented as the mean±SD.

As shown in the time-averaged velocity streamline maps (**Figure 7B**), compared with patients without aneurysms, those who developed aneurysms exhibited slight helical turbulence at the junction of the SA and the SMA, along with a more tortuous vascular course. Quantitative analysis revealed that, prior to aneurysm formation, patients who subsequently developed aneurysms already had significantly increased flow velocity and flow rate in the SA (velocity: *P*<0.001; flow rate: *P*=0.025, **Figure 7F,G**). However, analysis of local pressure isopleth distribution showed no statistically significant differences in mean SA pressure, whether overall, proximal, or distal, between the two groups (**Figure 7C,H**).

In addition, we calculated the TAWSS and OSI in the local region. The TAWSS contour maps are shown in **Figure 7D**. Quantitative analysis demonstrated that, compared with individuals with aberrant SA without aneurysms, those who subsequently developed aneurysms had lower TAWSS in both the proximal and distal segments of the SA (proximal: 5.7 vs. 5.3, *P*<0.001; distal: 23.9 vs. 11.5, *P*<0.001, **Figure 7I,J**). This indicates that, in the context of aberrant SA anatomy, individuals with lower TAWSS in the SA are more prone to developing SAAs. Precise quantitative analysis of OSI also revealed that, prior to aneurysm formation, the OSI at the junction of the SA and the SMA was already significantly elevated in individuals who subsequently developed aneurysms (*P*=0.017, **Figure 7K**).

In-depth analysis of the morphological characteristics of the restored pre-aneurysmal state revealed that the SA in these patients was generally more tortuous. Quantitative analysis of the tortuosity index further confirmed this observation (*P*<0.001, **Figure 7L**). Taken together with our previous morphological findings, we propose that unfavorable anatomical structure is the key determinant of whether an individual with aberrant SA anatomy will develop aneurysms.

### Predicting the occurrence of SAA in aberrant SA using machine learning

Given that the follow-up analysis of this study suggested that the occurrence of SAAs in individuals with aberrant SA anatomy may be highly correlated with the hemodynamic status induced by their morphological characteristics, we trained nine different ML models based on five morphological parameters (*C_dSA_*, *γ_SA-pA_*, *γ_SMA-aA_*, *C_pSA_/C_SMA_*, and *TI*) extracted from CTA 3D reconstruction images of 195 subjects, and evaluated their performance on a test dataset to predict the future risk of SAA development in patients with this anatomical variation.

**Figure 8A** presents the Pearson correlation heatmap among the five morphological parameters. The results showed no highly collinear feature pairs (|r| > 0.85) among the parameters, indicating good independence of the selected morphological features for direct use in subsequent modeling. To prevent overfitting, we performed dimensionality reduction on the features. The principal component analysis scree plot showed that the first two principal components cumulatively explained 55.33% of the total variance, and the first three components explained 74.76%. Based on the 90% variance explanation criterion, five principal components were retained (**Figure 8B1**). The scatter plot of the first two principal components (PC1 vs PC2) showed a certain separation trend between the two groups in the low-dimensional space, although some overlap remained, suggesting that the morphological parameters have moderate discriminative potential (**Figure 8B2**).

**Figure 8.**
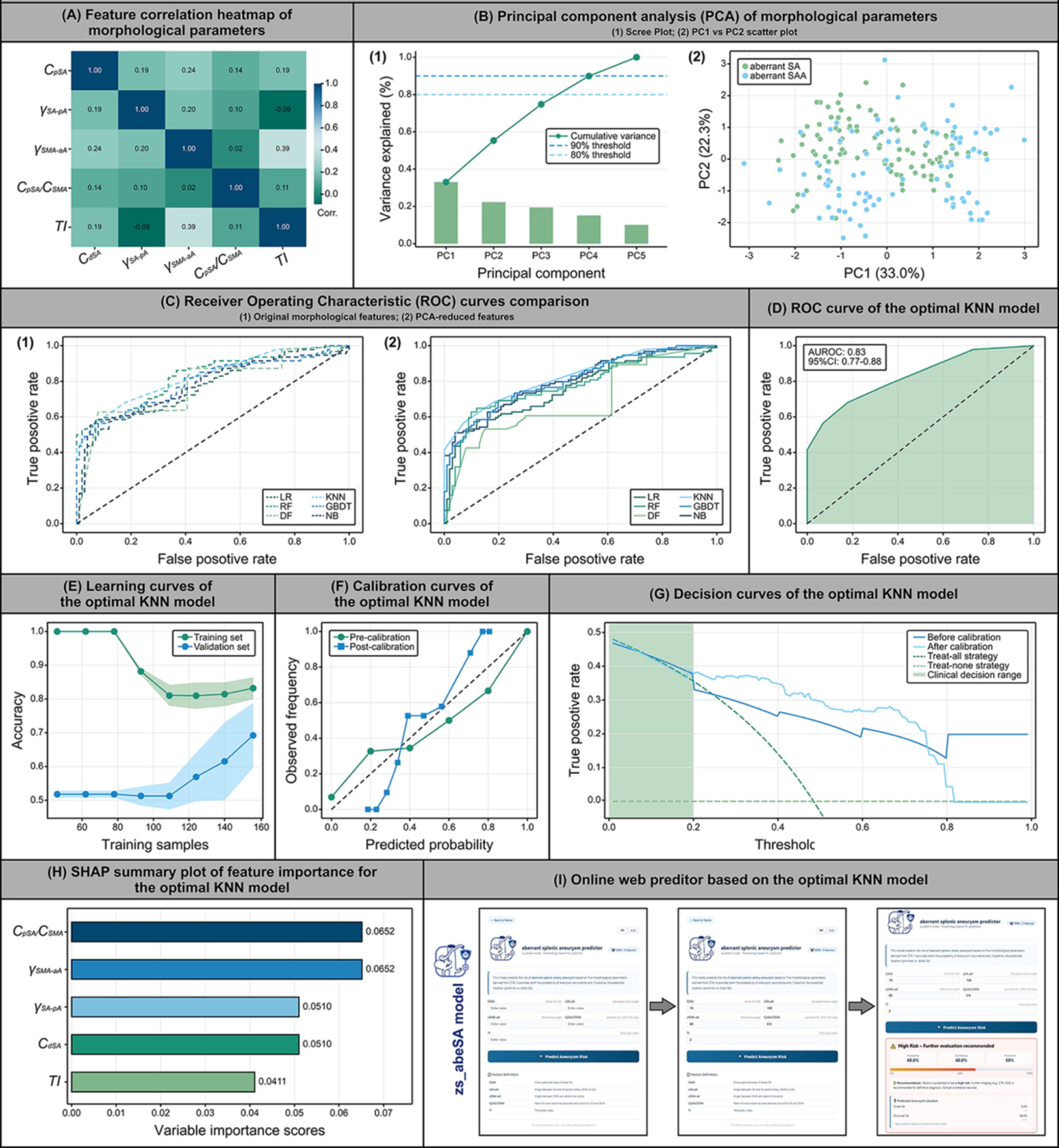
Development and evaluation of the zs_abeSA model. **(A)** Pearson correlation heatmap of the five morphological parameters. **(B1)** Scree plot of PCA. **(B2)** Scatter plot of the first two principal components. **(C1)** ROC curves of the top six ML models under leave-one-out cross-validation. **(C2)** ROC curves of the top six ML models after PCA dimensionality reduction. **(D)** ROC curve of the optimal KNN model with 95%CI (Bootstrap method) (AUROC=0.8277, 95%CI:0.770-0.878). **(E)** Learning curve of the KNN model (well-fitted, gap=0.140). **(F)** Calibration curve of the KNN model before and after Platt scaling (Brier score: pre-calibration 0.1662, post-calibration 0.1372; improvement: 0.0289). **(G)** Decision curve analysis. **(H)** SHAP feature importance ranking. **(I)** User interface of the zs_abeSA model web application. ML, machine learning. PCA, principal component analysis. ROC, receiver operating characteristic. SA, splenic artery.

Among the nine ML models, the KNN model achieved the best performance, with an area under the receiver operating characteristic curve (AUROC) of 0.8277 (95%CI: 0.770-0.878). Its secondary performance metrics were as follows: accuracy 0.754, specificity 0.822, sensitivity 0.681, negative predictive value 0.735, and positive predictive value 0.781 (**Table 5**). **Figure 8C1** shows the ROC curves of the top six performing ML models under leave-one-out cross-validation. **Figure 8C2** shows the ROC curves of each model after PCA dimensionality reduction; although the performance of some models varied slightly, KNN remained the best. **Figure 8D** presents the ROC curve of the optimal model (KNN) for predicting whether patients with aberrant SA anatomy will develop SAAs, with the shaded area indicating the 95%CI calculated by the Bootstrap method with 2,000 resampling iterations. The learning curve of the KNN model demonstrated that as the training sample size increased, the accuracy of both the training set and the validation set gradually converged, with a final gap of 0.14, indicating that the model did not overfit and had good generalization capability (**Figure 8E**). Calibration curves were used to assess the agreement between the model-predicted probabilities and the actual observed outcomes. After Platt scaling calibration, the model’s Brier score decreased from 0.166 before calibration to 0.137, an improvement of 0.029, suggesting excellent agreement between predicted and observed event probabilities (**Figure 8F**). Decision curve analysis was performed to evaluate the clinical net benefit of the KNN model. As shown in **Figure 8G**, within the threshold probability range of 0-0.5, the calibrated decision curve lay above both the treat-all and treat-none strategies, suggesting that using this model for clinical decision-making within this threshold range yields a positive net benefit and can serve as an auxiliary tool for clinical decision-making. Furthermore, we analyzed feature importance using the SHAP method. The results showed that *C_pSA_/C_SMA_* and *γ_SMA-aA_* were the two most important morphological parameters contributing to the prediction, with high values significantly associated with an increased predicted probability of aneurysm occurrence (**Figure 8H**). Finally, based on the above results, we developed an online prediction tool interface, termed the zs_abeSA model, using the optimal KNN model. This tool has been deployed on an Alibaba Cloud server. After users input the five morphological parameters, the system returns the occurrence probability and location prediction results in real time, demonstrating good clinical operability (**Figure 8I**). In conclusion, the zs_abeSA model is a high-performance and interpretable predictive tool for aberrant SAAs that can assist clinicians in developing personalized surveillance strategies in clinical practice, offering substantial clinical value and potential for further optimization.

**Table 5.**
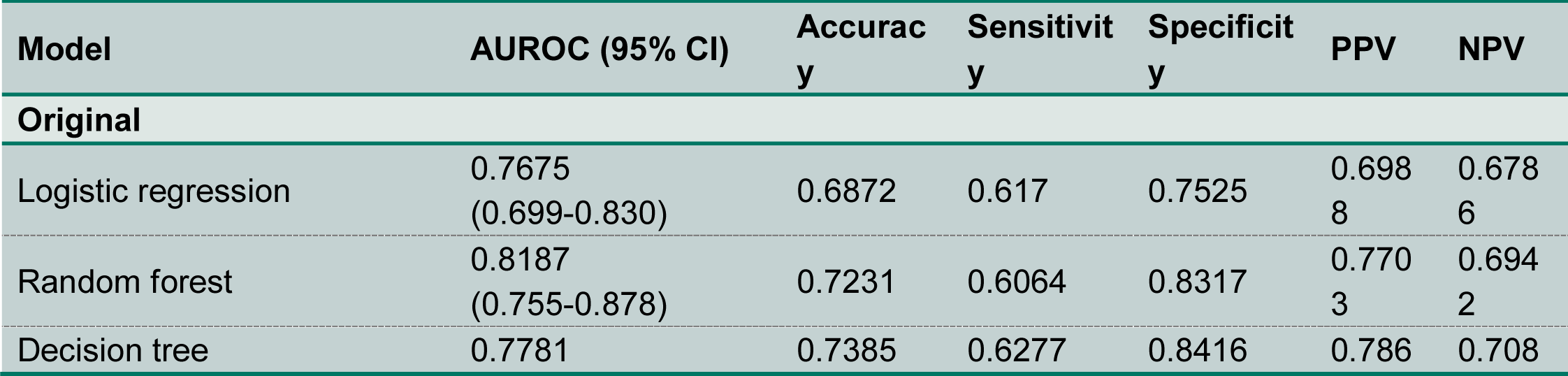
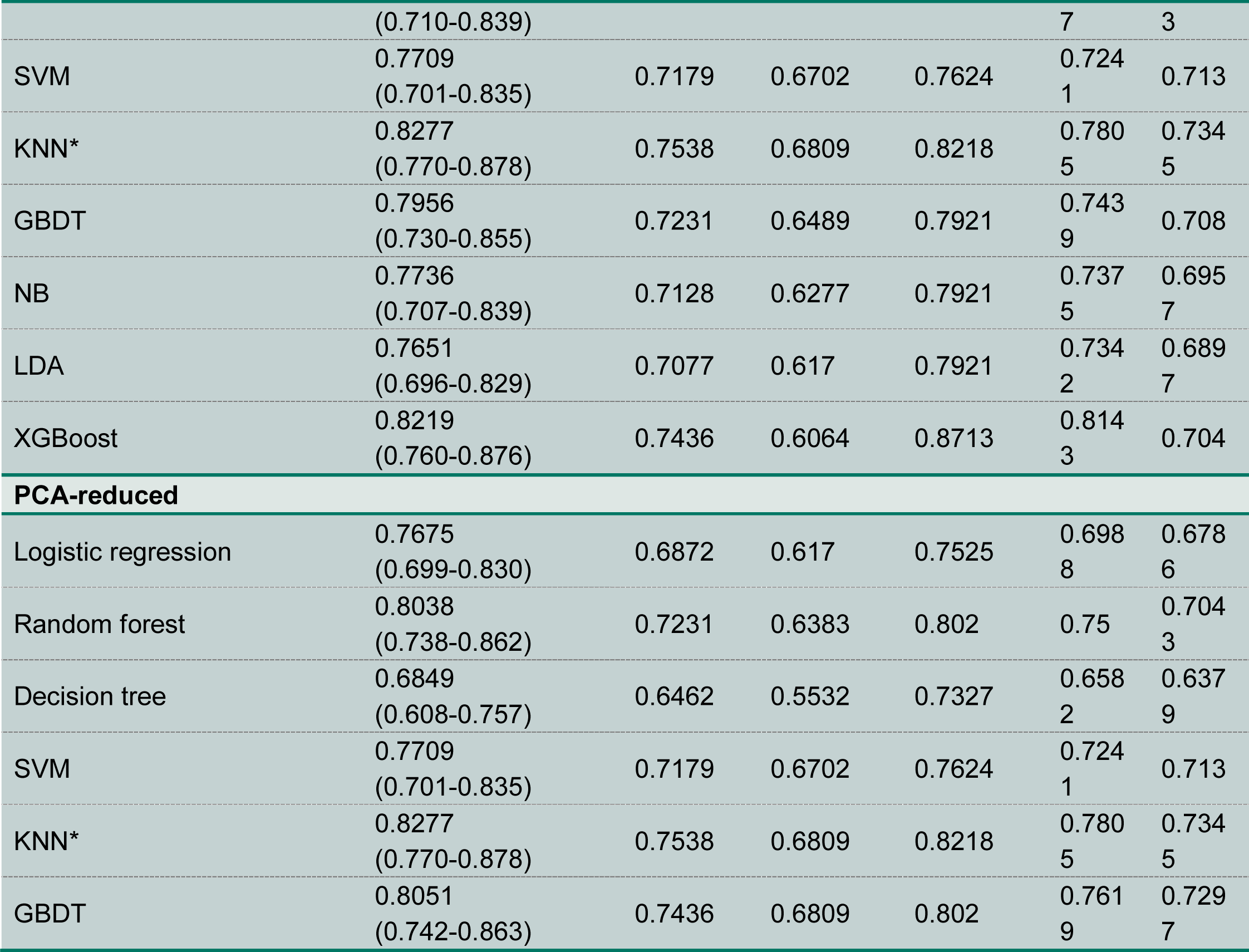
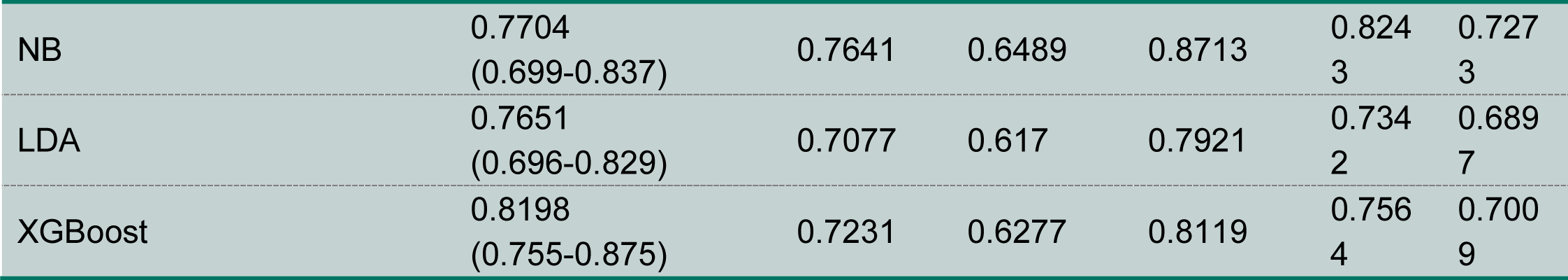
Model performance on testing data for prediction the occurrence of SAA in aberrant SA. AUROC, area under the receiver operating characteristic curve. CI, confidence interval. GBDT, gradient boosting decision tree. KNN, K-Nearest neighbors. NB, Naive Bayesian. NPV, negative predictive value. PPV, positive predictive value. SA, splenic artery. SAA, splenic artery aneurysm. SVM, support vecto

## Discussion

Aberrant SA is a rare anatomical variant in which the SA arises from the SMA rather than the CA, with a reported prevalence of less than 1% in the general population [34]. A systematic review encompassing 3,132 specimens reported that the proportion originating from this site was approximately 0.7% [35]. As a rare anatomical anomaly, it has long lacked large-scale cohort studies, and the understanding of its potential pathogenic implications remains limited. Our clinical experience suggests that patients with this anatomical variant are highly prone to developing SAAs; however, no mechanistic studies to date have elucidated why this anatomical anomaly predisposes to the high incidence of local SAAs.

This study enrolled 195 patients with aberrant SA anatomy from 16 centers across China between 2008 and 2026, representing the largest known cohort of this rare anatomical variant worldwide. The cohort included 94 patients with concomitant SAAs and 101 patients without aneurysms. Based on these data, the estimated incidence of SAAs in the aberrant SA population was approximately 48.2%. However, this figure should be interpreted with caution, as it may be subject to selection bias-asymptomatic individuals or those without aneurysms are less likely to undergo CTA examination, and even when examined, cases may not have been captured if the report did not explicitly document the anatomical variation. In addition, to facilitate comparative analysis, we also included a cohort of 100 patients with normal anatomy and a cohort of 100 patients with SAAs in the setting of normal anatomy as control groups. This large-scale, multicenter case series enabled us to move beyond the limitations of previous case reports and provide the first systematic characterization of the morphological spectrum of this anatomical variant, as well as to establish a comprehensive model linking morphology, hemodynamics, and aneurysm development. The increased statistical power afforded by the large sample size supported robust subgroup analyses and multivariable adjustments, while also enabling the development and internal validation of a predictive model, thereby substantially enhancing the generalizability and clinical utility of our findings.

Based on 3D reconstruction of abdominal CTA images from this multicenter retrospective cohort, we collected morphological data and performed hemodynamic analyses of the abdominal vasculature for each patient, and further explored the potential mechanisms underlying aneurysm development in the context of aberrant SA anatomy. We found that aberrant SAAs were significantly larger than common SAAs and predominantly located in the proximal segment of the SA (86.4%). In contrast, common SAAs were mainly located in the middle-distal segment or at the splenic hilum, with approximately 74%-87% of cases occurring in the distal third of the SA [36,37]. The distinctly opposite distribution patterns strongly suggest that the pathogenesis of aberrant SAAs differs from that of common SAAs, and that the former may carry a higher potential risk of rupture and a more aggressive clinical course. Common SAAs are generally attributed to atherosclerosis, medial fibromuscular dysplasia, or portal hypertension, which are conditions that tend to affect distal arteries. In contrast, the proximal predilection of aberrant SAAs points toward a hemodynamically driven mechanism, wherein the anomalous origin from the SMA exposes the proximal SA to abnormal flow conditions. Our study demonstrated that the aberrant SA exhibited a smaller branching angle with its parent vessel, a larger cross-sectional area ratio, and a more tortuous course. These unfavorable morphological features predispose the aberrant SA-particularly its proximal lateral wall-to receive greater blood flow and bear higher pressure. Hemodynamic analysis further corroborated the morphological findings, revealing that the SA arising from the SMA is subjected to a fundamentally different hemodynamic environment compared with that arising from the CA. CFD analysis showed that the TAWSS at the proximal segment of the aberrant SA was significantly lower than that of the normal SA. Low WSS is a well-established pro-atherogenic and pro-aneurysmal stimulus that induces endothelial dysfunction, upregulates inflammatory mediators, and promotes matrix metalloproteinase activity, ultimately leading to progressive wall weakening and aneurysmal dilatation. The chronically low-shear environment at the proximal aberrant SA provides a direct mechanistic explanation for the proximal predilection of these aneurysms. Furthermore, the OSI at the SA-parent vessel junction was significantly elevated and was inversely correlated with the branching angle. High OSI reflects marked directional changes in blood flow during the cardiac cycle, imposing mechanical perturbation on the endothelium at the bifurcation apex. The hemodynamic profile of this region-low TAWSS, high OSI, and increased flow burden-constitutes a ‘perfect storm’ for aneurysm initiation, explaining both the higher prevalence and the larger size of aneurysms in this population. Through the above analyses, we have elucidated the morphological characteristics and hemodynamic remodeling that predispose individuals with aberrant SA anatomy to aneurysm development, and have delineated the mechanistic chain by which these factors promote the occurrence and progression of SAAs. These findings provide a theoretical basis for predicting the occurrence of aberrant SAAs and may also offer insights for mechanistic studies of other VAAs and/or intracranial aneurysms.

A clinically important question addressed in this study is: among individuals with the same anatomical variant, why do some develop aneurysms while others do not? We first followed a small cohort of non-aneurysmal patients with aberrant SA anatomy (n=14, mean follow-up 36.8±22.1 months, mean 2.9±0.8 visits), and found that vessel dimensions remained stable during follow-up, with no significant changes in either proximal or distal diameters. This suggests that aneurysm development in this population is not a chronic degenerative process, but rather an acute or subacute event triggered by adverse morphological and hemodynamic conditions. However, the limited sample size of this follow-up cohort, together with the lack of follow-up data from patients with aberrant SAAs (who almost invariably undergo surgical treatment shortly after diagnosis), underscores the need for future studies with larger cohorts and longer-term follow-up. To further explore the underlying causes of individual differences, we performed a cross-sectional comparative analysis between non-aneurysmal and aneurysmal aberrant SA patients, using pre-aneurysmal morphological restoration for the latter. The results showed no significant differences in proximal or distal pressures between the two groups. However, patients who subsequently developed aneurysms had significantly lower distal TAWSS in their restored pre-aneurysmal state compared with those who remained aneurysm-free (4.2Pa vs. 3.6Pa, *P*<0.0001). This finding suggests that even within the same anatomical variant, subtle individual variations in vessel geometry can influence the local hemodynamic environment (specifically, the spatial distribution pattern of WSS rather than pressure elevation per se), thereby determining susceptibility to aneurysm formation. Thus, aneurysm development in the aberrant SA is not an inevitable consequence of the anatomical variant, but rather a probabilistic event modulated by individual geometric and hemodynamic factors. This aligns with our clinical observation that some patients may develop VAAs (including SAAs) over relatively short time intervals, particularly during pregnancy or under acute hemodynamic stress. For patients incidentally found to have aberrant SA anatomy without aneurysms, aggressive imaging surveillance may not be necessary; instead, individualized risk assessment is recommended. To address the aforementioned clinical need for individualized risk assessment in patients with aberrant SA anatomy, we developed and validated the zs_abeSA predictive model and encapsulated it as an interactive web application to facilitate its clinical translation and application. This model can predict both the probability and the potential location of future SAA development in individuals with aberrant SA anatomy. The model achieved an AUROC of 0.828, indicating good discriminative ability for identifying individuals at risk of aneurysm development. The model was trained using morphological parameters that directly influence the hemodynamic environment, achieving favorable predictive performance. This finding is consistent with our mechanistic observations and further supports the notion that morphology and hemodynamics are inextricably linked, and that morphological parameters can serve as reliable surrogates for hemodynamic risk. Among these parameters, the *C_pSA_*/*C_SMA_* ratio is particularly noteworthy as one of the most important predictors. This ratio reflects the relative caliber of the SA compared with its parent vessel, directly determining the flow distribution. A higher ratio indicates a larger SA relative to the SMA, allowing a greater proportion of SMA flow to enter the SA, thereby increasing WSS and pressure burden on the proximal vessel wall. This parameter can be derived directly from routine CTA without the need for complex CFD modeling, making it a practical and easily accessible bedside risk stratification tool. We deliberately adopted a parsimonious modeling strategy. Although incorporating additional parameters might yield marginal statistical gains in larger cohorts, our cohort of 195 patients already represents the largest known study population in this field. The use of five carefully selected morphological features helps prevent overfitting and ensures that the zs_abeSA model remains both cost-effective and clinically accessible, requiring only standard CTA measurements without the need for complex CFD simulations.

From a clinical perspective, the model’s moderate sensitivity (0.681) and specificity (0.822) require careful interpretation. The higher specificity suggests that the model is more reliable in ruling out disease, which may be particularly useful for screening to avoid unnecessary follow-up in a rare condition. However, the relatively lower sensitivity implies a non-negligible risk of false negatives, which, in the context of a potentially life-threatening condition, underscores the importance of using the model as an adjunct to clinical judgment, not as a replacement. Future prospective studies are needed to better characterize the real-world impact of misclassification. Based on the above findings, we propose the following recommendations for the clinical management of patients with aberrant SA anatomy:

**1) Screening recommendations.** For patients in whom aberrant SA anatomy is incidentally detected on CTA, dedicated evaluation for SAAs is recommended, with particular attention to the proximal SA. Given that the prevalence of aneurysms in this population is substantially higher than that in the general population, targeted screening is justified.
**2) Individualized surveillance intervals.** For patients without aneurysms, the longitudinal follow-up in this study demonstrated stable vessel dimensions, suggesting that annual surveillance may not be necessary. Nevertheless, baseline imaging followed by risk stratification using the zs_abeSA model is recommended. Patients identified as high-risk by the model may benefit from closer surveillance, whereas low-risk patients may be followed up with less frequent imaging.
**3) Lower threshold for intervention.** The larger aneurysm size, proximity to major vessels, and more aggressive hemodynamic profile of aberrant SAAs may support a lower intervention threshold compared with common SAAs. While the conventional intervention threshold for common SAAs is generally 2cm, our findings suggest that for aberrant SAAs (particularly in women of childbearing age, who face an increased risk of rupture during pregnancy, or in patients identified as having high hemodynamic risk by the model), intervention may be considered at smaller sizes.
**4) Surgical planning considerations.** The predilection of aberrant SAAs for the proximal segment, close to the origin of the SMA, necessitates special attention to preserving SMA flow during endovascular interventions. The planning of surgical or endovascular repair should fully account for hemodynamic alterations in the parent vessel to avoid unintended consequences on distal perfusion resulting from disruption of the abnormal flow pattern.

## Conclusion

In conclusion, this study, based on data from 195 patients across 16 centers, demonstrates that aberrant SA anatomy, defined as the SA arising from the SMA, is a significant independent risk factor for SAA formation. The pathogenic mechanism involves a cascade from anatomical variation to morphological remodeling, including smaller branching angle, increased cross-sectional area ratio, and increased tortuosity index, then to hemodynamic derangement, including low proximal TAWSS, high OSI, and increased flow burden, ultimately leading to aneurysm formation. Aberrant SAAs are larger in size and predominantly located in the proximal SA. The stability of vessel dimensions observed during follow-up in non-aneurysmal individuals with aberrant SA anatomy suggests that aneurysm development is not an inevitable consequence of the anatomical variant, but rather a probabilistic event determined by individual geometric and hemodynamic factors. The zs_abeSA predictive model provides a practical tool for risk stratification and clinical decision-making in aberrant SAAs. These findings have direct clinical implications for screening, surveillance, and treatment strategies in patients with this anatomical variant, and provide a theoretical framework for integrating morphological and hemodynamic assessment into the clinical management of VAAs.

### Study limitation

Several limitations of this study should be acknowledged when interpreting the results.

First, this study was a retrospective cohort design, which is inherently subject to selection bias. The inclusion of patients based on available CTA data may have introduced screening bias, as patients with more severe symptoms or higher clinical suspicion were more likely to undergo imaging, whereas asymptomatic individuals or those without overt disease were less likely to receive CTA examination, or even if examined, may have been missed if the report did not explicitly document the anatomical variation. Although we attempted to mitigate this through matching with control cohorts, residual confounding cannot be entirely excluded.

Second, the sample size, particularly for the CFD analysis and longitudinal follow-up, was relatively limited. The rarity of aberrant SA anatomy precluded a larger sample. Although this study represents the largest cohort of aberrant SAAs to date, some subgroup analyses may have been underpowered to detect smaller but clinically meaningful differences. Future multicenter studies with larger cohorts are warranted to validate our findings.

Third, the CFD simulations were based on several simplifying assumptions, including the treatment of blood as a Newtonian fluid and the assumption of rigid vessel walls. While these assumptions are standard practice in large-vessel hemodynamic analyses, they may not fully capture the complexity of in vivo flow, including the effects of vessel wall compliance and non-Newtonian blood behavior. Future studies employing fluid-structure interaction models may provide more detailed insights.

Fourth, although our predictive model demonstrated good performance, it has not been prospectively validated. Given the rarity of this disease, large-scale prospective deployment and validation are not feasible. The generalizability of the zs_abeSA model to other populations and imaging protocols remains to be established.

Fifth, our study focused on morphological and hemodynamic factors and did not incorporate genetic, biochemical, or inflammatory biomarkers. Given the growing evidence of the role of inflammation and matrix remodeling in aneurysm pathogenesis, future studies should integrate circulating biomarkers or tissue-level analyses to provide a more comprehensive understanding of the disease process.

Sixth, although clinical follow-up data were available for a subset of patients, the follow-up duration was relatively short. Long-term follow-up beyond 5 years is needed to determine the ultimate fate of non-aneurysmal aberrant SA and the lifetime risk of aneurysm formation.

### Machine Learning Checklist

The ML modeling process in this study adhered to the recommended standards outlined in the Proposed Requirements for Cardiovascular Imaging-Related Machine Learning Evaluation (PRIME) checklist. We have reported the study design objectives, data preprocessing and standardization methods, feature selection strategy, model selection and hyperparameter optimization process, model evaluation metrics (including AUROC, accuracy, sensitivity, specificity, positive predictive value, and negative predictive value), as well as calibration curve and decision curve analysis results. Model reproducibility is ensured through open-source code (GitHub), and limitations are addressed in the Discussion section. The completed PRIME checklist is provided below.

| **PRIME Checklist for Standardized Reporting of Machine Learning Investigations** |  |  |
| --- | --- | --- |
| Section | Checklist item | Response / Location in Manuscript |
| **1. Designing the Study Plan** |  |  |
| 1.1 | Describe the need for the application of machine learning to the dataset | Introduction, page 7 - 'To date, no ML-based predictive model has been specifically developed for SAAs, particularly in the context of aberrant SA anatomy. Our study addresses this gap by developing and validating a two-stage ML model that predicts both the risk of occurrence and the potential location of aneurysms in patients with this rare anatomical variant.' |
| 1.2 | Describe the objectives of the machine learning analysis | Methods, page 15 - 'This study employed a two-stage prediction strategy to jointly model the prediction of SAA occurrence and localization, simulating the real-world clinical decision-making process: first determining whether a patient has an aneurysm, and if so, further identifying its specific location.' |
| 1.3 | Define the study plan | Methods, page 15 - 'Based on five morphological parameters, nine ML algorithms were incorporated to construct binary classification models for SAA prediction (occurrence vs. non-occurrence).' |
| 1.4 | Describe the summary statistics of baseline data | Results, page 20; Table 3 - Baseline characteristics of all study participants (n=395), including demographics, comorbidities, and clinical features across four groups. |
| 1.5 | Describe the overall steps of the machine learning workflow | Methods, page 9-10; Figure 1B - Workflow includes: (1) data preprocessing and normalization, (2) feature selection, (3) model training with LOOCV, (4) model evaluation, and (5) model deployment as a web application. |
| **2. Data Standardization, Feature Engineering, and Learning** |  |  |
| 2.1 | Describe how the data were processed in order to make it clean, uniform, and consistent | Methods, page 16 - 'All features were standardized using Z-score normalization to eliminate the effects of different scales.' |
| 2.2 | Describe whether variables were normalized and if so, how this was done | Methods, page 16 - 'All features were standardized using Z-score normalization to eliminate the effects of different scales.' |
| 2.3 | Provide details on the fraction of missing values (if any) and imputation methods | Methods, page 15 - 'No missing values were present in the morphological parameters used for model training.' |
| 2.4 | Describe any feature selection processes applied | Methods, page 15 - 'Although a total of 12 morphological parameters were initially measured, we subsequently refined the feature set based on multicollinearity diagnostics (variance inflation factor <5) and clinical interpretability.' |
| 2.5 | Identify and describe the process to handle outliers if any | Methods, page 15 - 'No outliers were identified or removed; all samples were retained for model training.' |
| 2.6 | Describe whether class imbalance existed, and which method was applied to deal with it | Methods, page 16 - 'Preliminary analysis revealed that the dataset comprised 94 cases (48.2%) with aneurysms and 101 cases (51.8%) without aneurysms, indicating a relatively balanced class distribution; therefore, no oversampling techniques were applied.' |
| **3. Selection of Machine Learning Models** |  |  |
| 3.1 | Explicitly define the goal of the analysis e.g., regression, classification, clustering | Methods, page 15 - 'Nine ML algorithms were incorporated to construct binary classification models for SAA prediction (occurrence vs. non-occurrence)." |
| 3.2 | Identify the proper learning method used (e.g., supervised, reinforcement learning etc.) to address the problem | Methods, page 15 - 'Supervised learning was employed for both the occurrence prediction and location prediction tasks.' |
| 3.3 | Provide explicit details on the use of simpler, complex, or ensemble models | Methods, page 15-16 - 'nine ML algorithms were incorporated to construct binary classification models for SAA prediction (occurrence vs. non-occurrence), including logistic regression, random forest, SVM, KNN, decision tree, GBDT, Naïve Bayes, LDA, and XGBoost.' |
| 3.4 | Provide the comparison of complex models against simpler models if possible | Results, page 30; Figure 8C1; Table 5 - 'Among the nine models compared, KNN achieved the best performance (AUROC = 0.8277), outperforming logistic regression (AUROC = 0.716) and other complex models.' |
| 3.5 | Define ensemble methods, if used | Methods, page 8 - Random forest, GBDT, and XGBoost are ensemble methods that combine multiple weak learners to improve predictive performance. |
| 3.6 | Provide details on whether the model is interpretable | Methods, page 17 - 'Feature importance was quantified using SHAP (SHapley Additive exPlanations) to enhance model interpretability.' |
| **4. Model Assessment** |  |  |
| 4.1 | Provide a clear description of data used for training, validation, and testing | Methods, page 16 - 'Leave-one-out cross-validation (LOOCV) was used for model evaluation. At each iteration, n-1 samples were used for training and 1 sample for validation.' |
| 4.2 | Describe how the model parameters were optimized (e.g., optimization technique, number of model parameters etc.) | Methods, page 8 - No systematic hyperparameter tuning was performed; default parameters were used for all models. The KNN model's performance was evaluated using LOOCV with default settings. |
| **5. Model Evaluation** |  |  |
| 5.1 | Provide the metric(s) used to evaluate the performance of the model | Results, page 30; Table 5 - Model performance was assessed using AUROC, accuracy, sensitivity, specificity, PPV, NPV, Brier score, calibration curves, and decision curve analysis. |
| 5.2 | Define the prevalence of disease and the choice of the scoring rule used | Methods, page 8; Results, page 20 - The prevalence of splenic artery aneurysms in the aberrant splenic artery population was 48.2% (94/195). The Brier score was used as the scoring rule to evaluate the calibration of predicted probabilities. |
| 5.3 | Report any methods used to balance the numbers of subjects in each class | Methods, page 16 - 'No oversampling techniques were applied, as the class distribution was relatively balanced (48.2% vs. 51.8%).' |
| 5.4 | Discuss the risk associated to misclassification | Discussion, page 37 - The model's high sensitivity (0.681) minimizes the risk of missing true positive cases, while the moderate specificity ensures that false positives are limited, balancing clinical utility with resource allocation. |
| **6. Best Practices for Model Replicability** |  |  |
| 6.1 | Consider sharing code or scripts on a public repository with appropriate copyright protection steps for further development and non-commercial use | Methods, page 19 - 'The source code and model files have been open-sourced on GitHub (https://github.com/RanXu1995/zs_abeSA-model).' |
| 6.2 | Release a data dictionary with appropriate explanation of the variables | Table 2; Figure 2 - A data dictionary defining all five morphological parameters ($C_{pSA}$, $\gamma_{SA-pA}$, $\gamma_{SMA-aA}$, $C_{pSA}/C_{SMA}$, $TI$) is provided in the |
|  |  | supplementary materials. |
| 6.3 | Document the version of all software and external libraries used | Methods, page 17 - 'All statistical analyses and ML modeling were performed using Python 3.14, with core libraries including scikit-learn, SHAP, and Matplotlib.' |
| **7. Reporting Limitations, Biases and Alternatives** |  |  |
| 7.1 | Identify and report the relevant model assumptions and findings | Discussion, page 39-40 - Several limitations should be acknowledged: retrospective design, relatively limited sample size for CFD analysis, and the model has not been prospectively validated. |
| 7.2 | If well performing models were tested on a hold-out validation dataset, detail the data of that validation set with the same rigor as that of training dataset | Methods, page 16 - 'LOOCV was used, ensuring that each validation sample was independent of the training set and drawn from the same distribution.' |

#### TRUE-AIM report card for Artificial Intelligence (AI) research

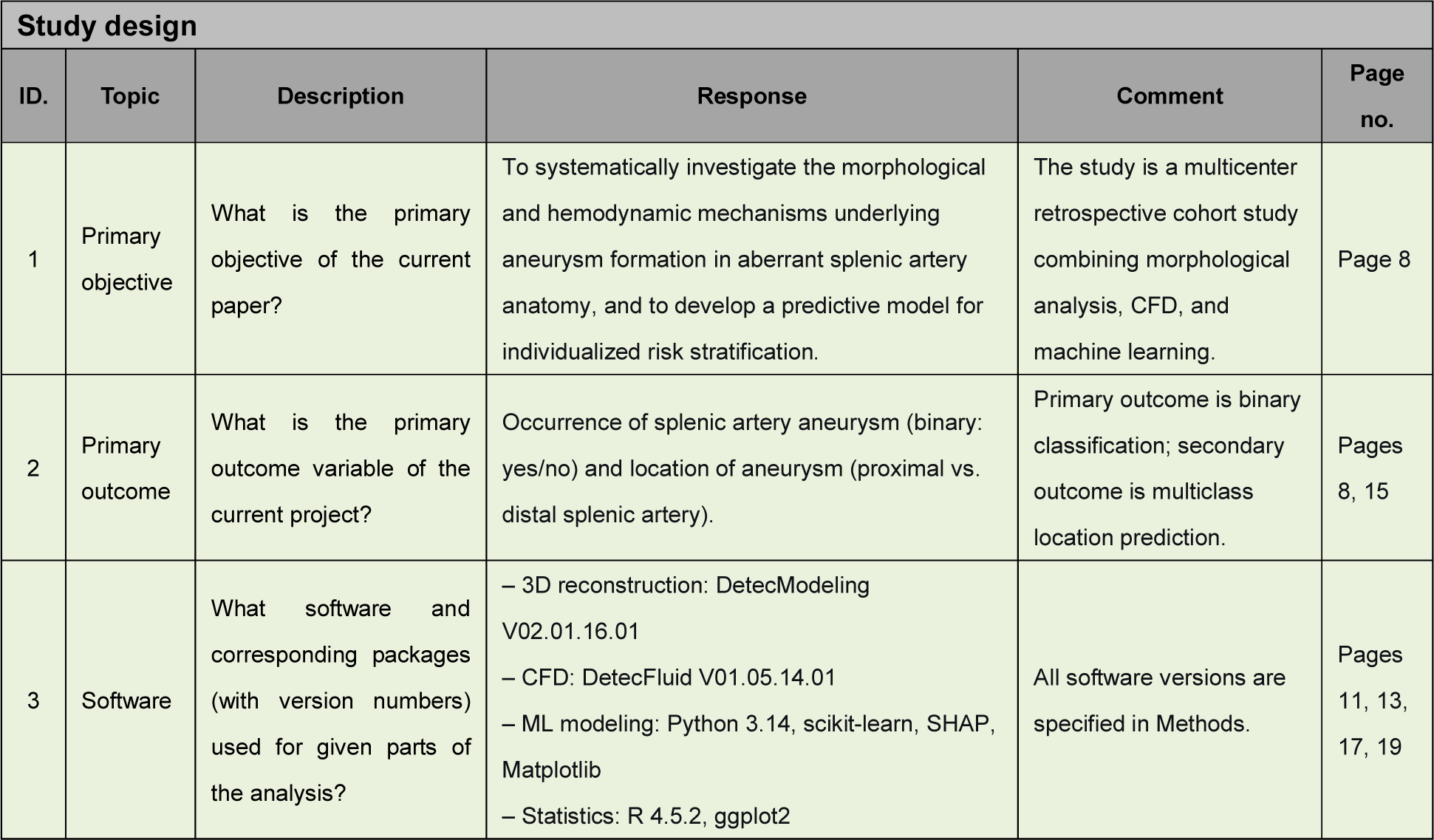
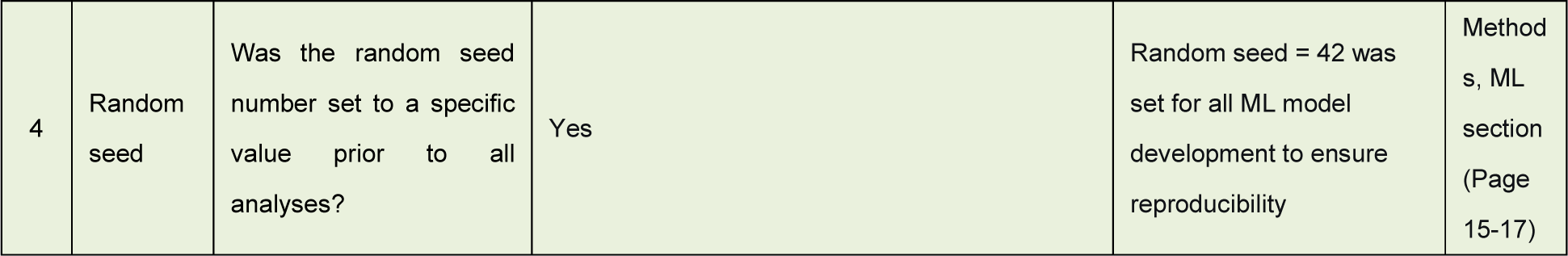

| Study design |  |  |  |  |  |
| --- | --- | --- | --- | --- | --- |
| ID. | Topic | Description | Response | Comment | Page no. |
| 1 | Primary objective | What is the primary objective of the current paper? | To systematically investigate the morphological and hemodynamic mechanisms underlying aneurysm formation in aberrant splenic artery anatomy, and to develop a predictive model for individualized risk stratification. | The study is a multicenter retrospective cohort study combining morphological analysis, CFD, and machine learning. | Page 8 |
| 2 | Primary outcome | What is the primary outcome variable of the current project? | Occurrence of splenic artery aneurysm (binary: yes/no) and location of aneurysm (proximal vs. distal splenic artery). | Primary outcome is binary classification; secondary outcome is multiclass location prediction. | Pages 8, 15 |
| 3 | Software | What software and corresponding packages (with version numbers) used for given parts of the analysis? | – 3D reconstruction: DetecModeling V02.01.16.01<br>– CFD: DetecFluid V01.05.14.01<br>– ML modeling: Python 3.14, scikit-learn, SHAP, Matplotlib<br>– Statistics: R 4.5.2, ggplot2 | All software versions are specified in Methods. | Pages 11, 13, 17, 19 |
| 4 | Random seed | Was the random seed number set to a specific value prior to all analyses? | Yes | Random seed = 42 was set for all ML model development to ensure reproducibility | Methods, ML section (Page 15-17) |

#### To be completed for all manuscripts that use AI as part of the Methods

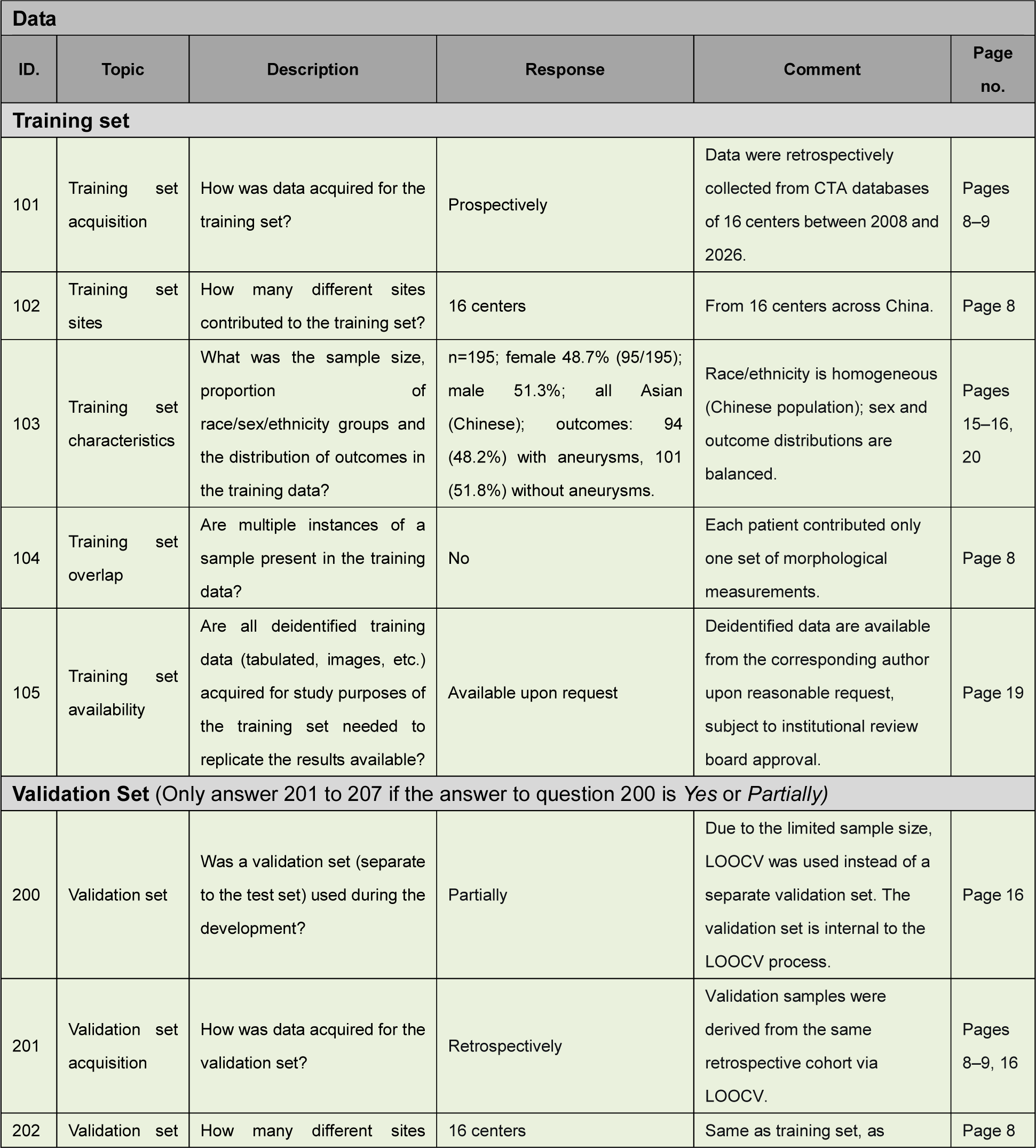
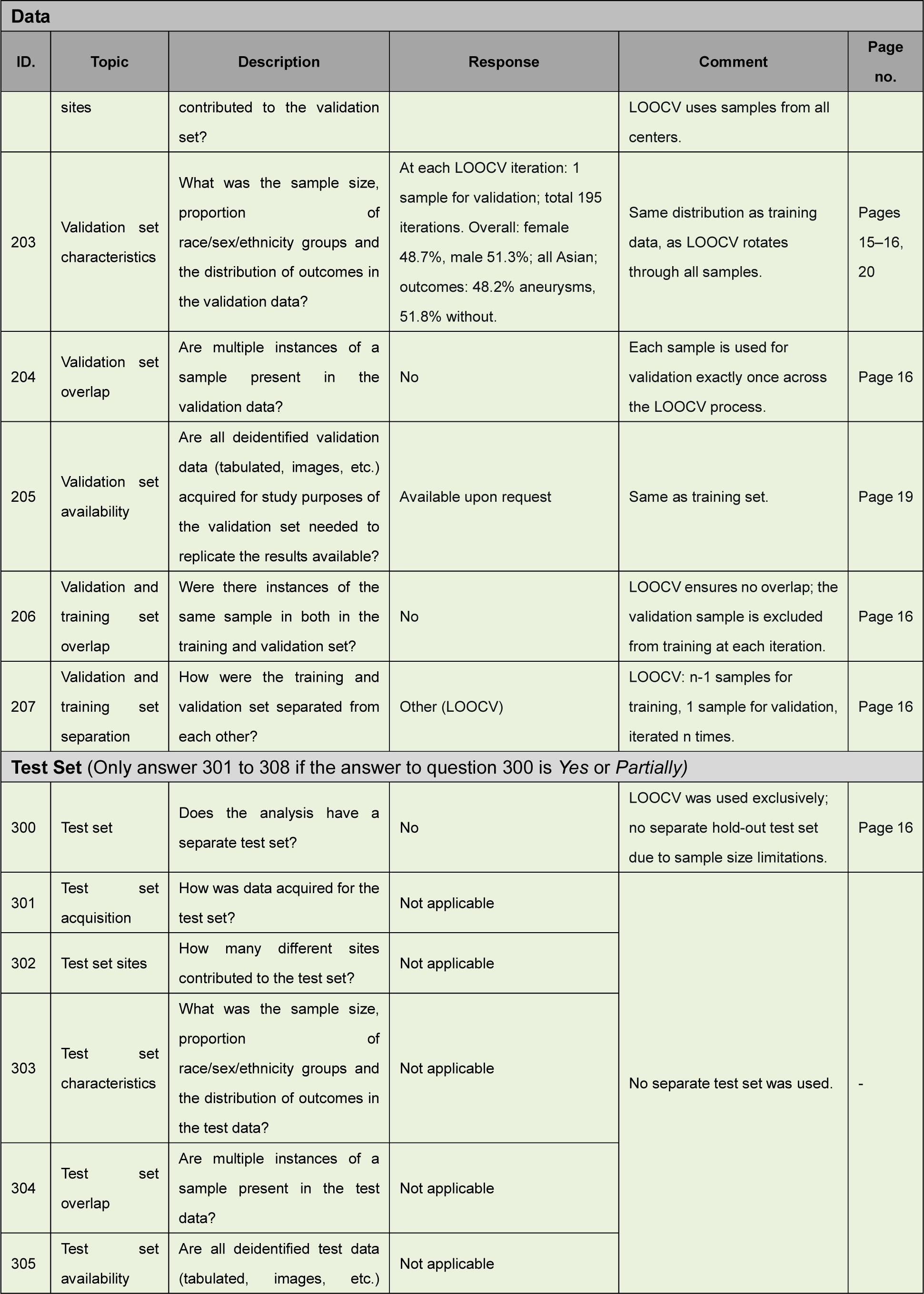
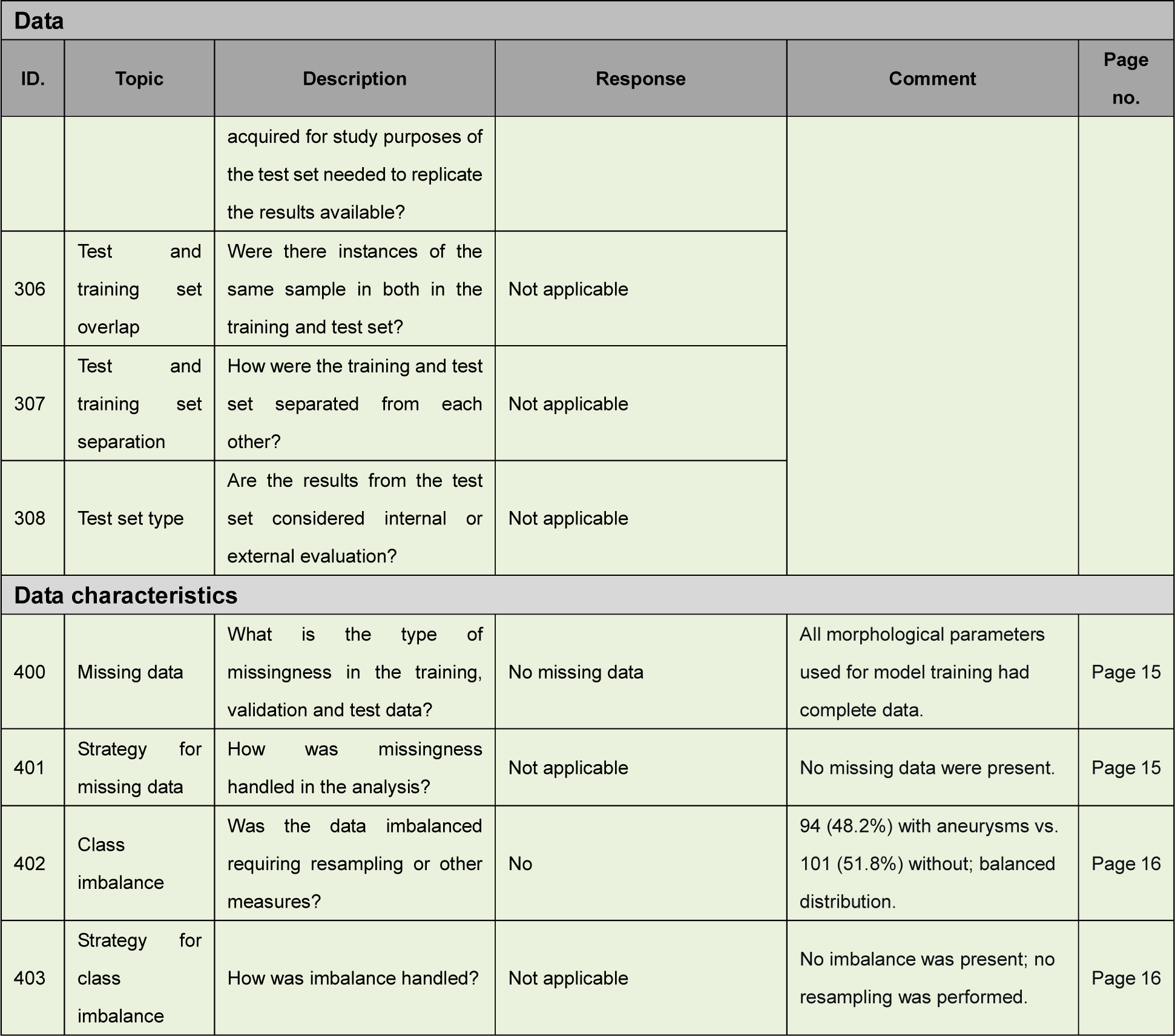

#### To be completed if applicable to the manuscript

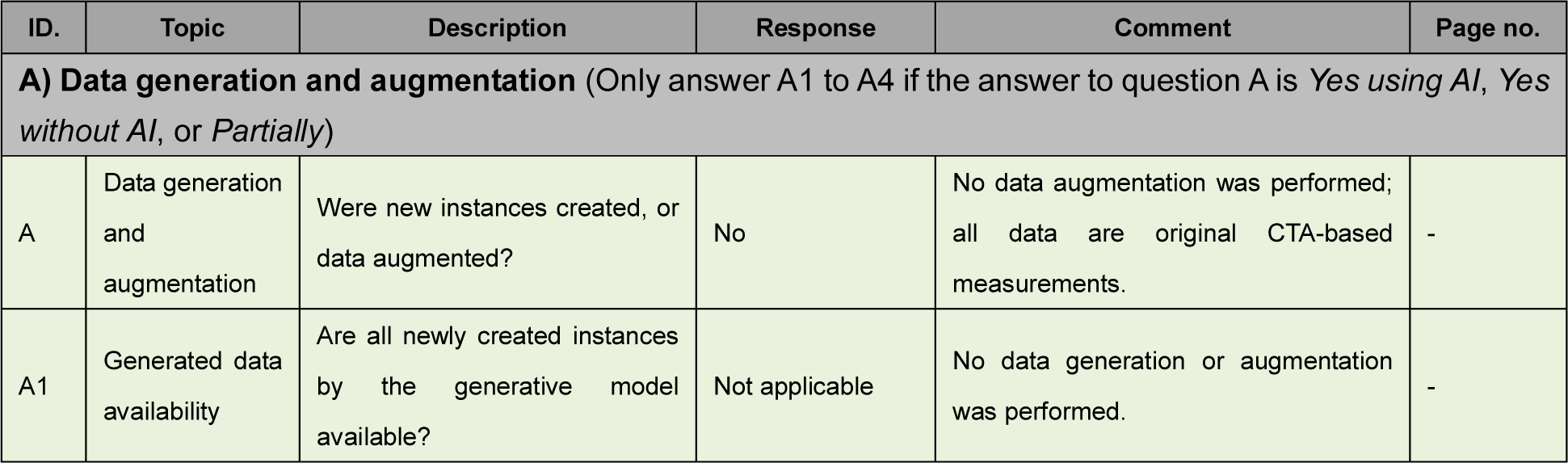
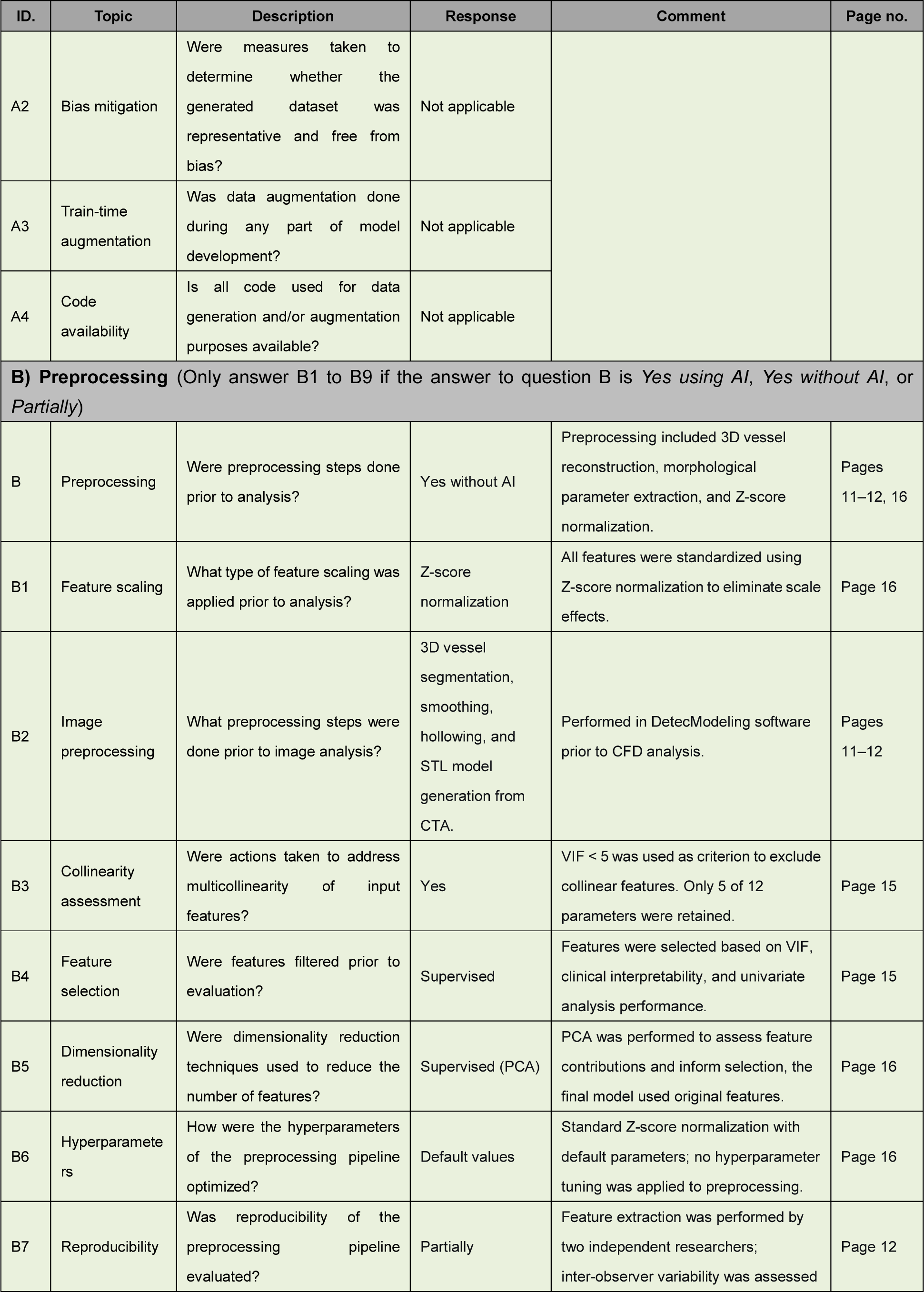
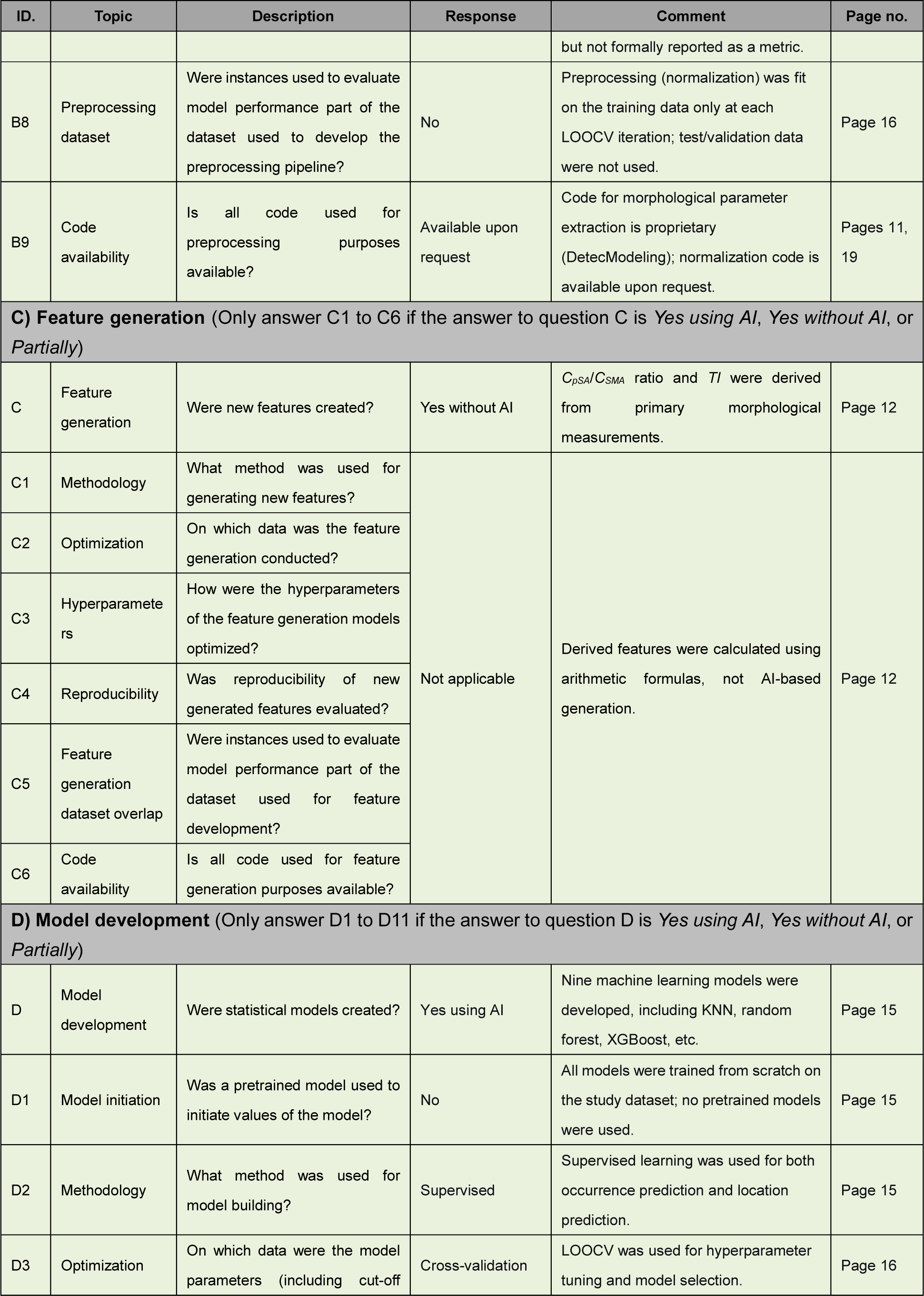
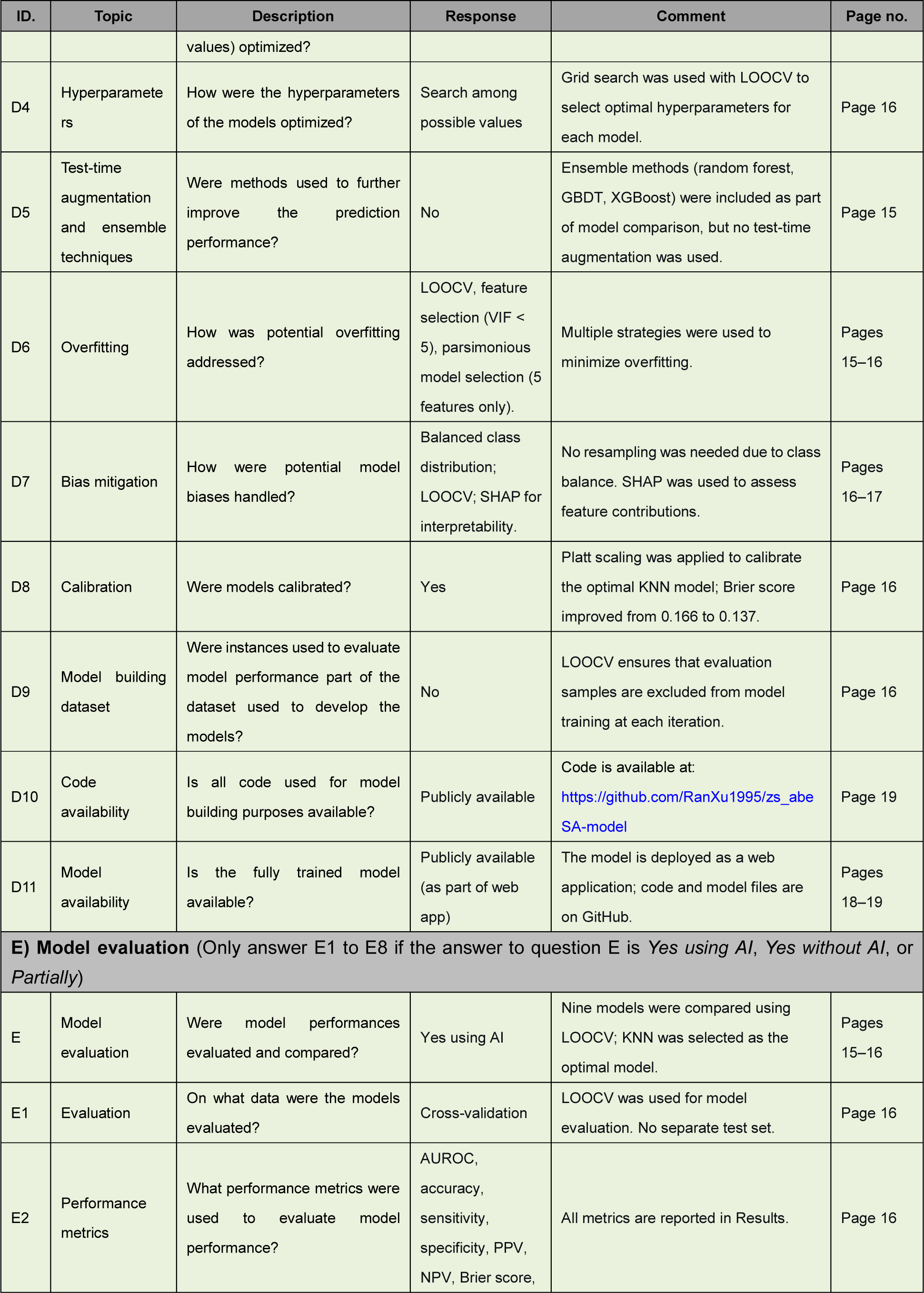
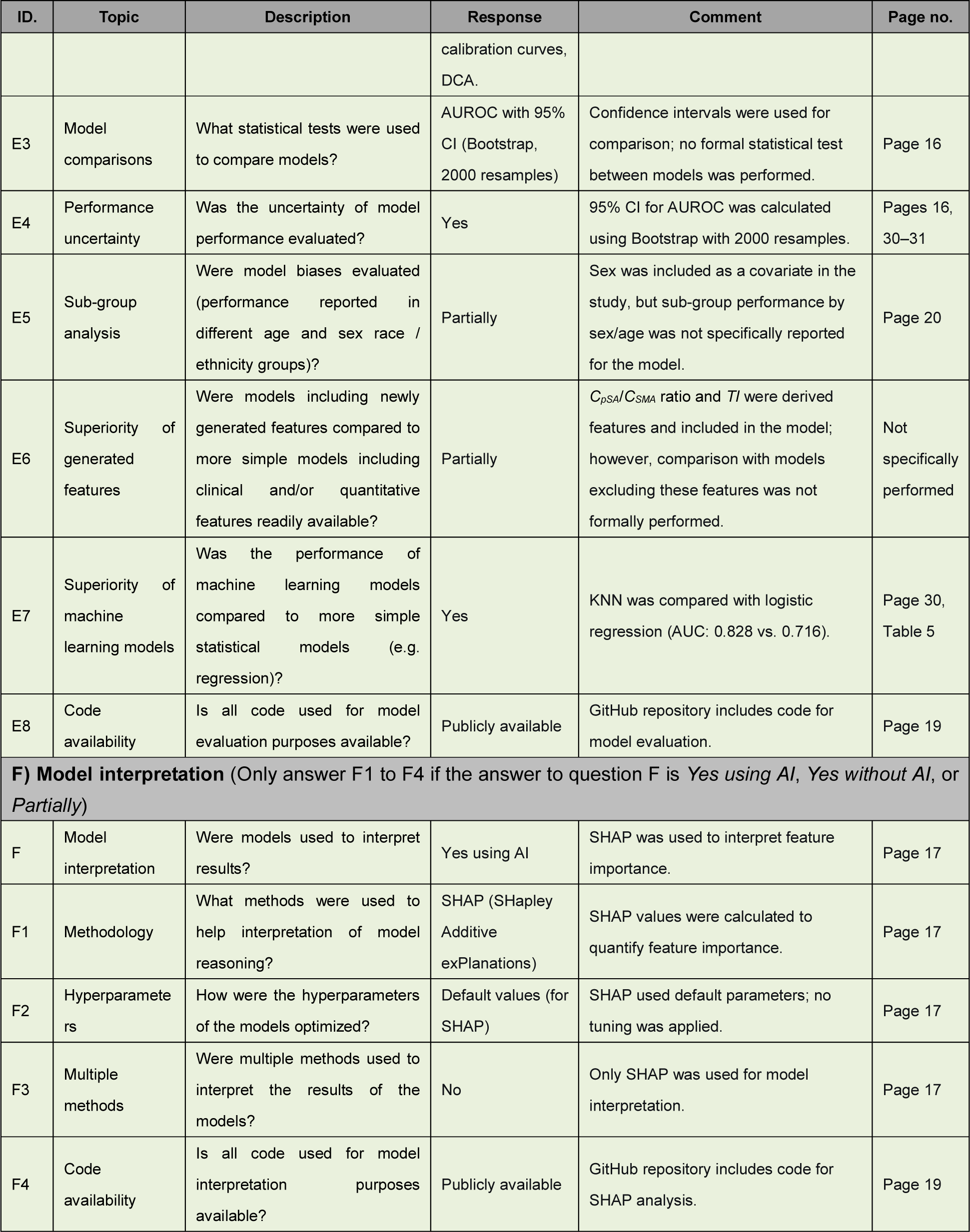

### Clinical Perspectives

#### Core clinical competencies

This study addresses the diagnostic and therapeutic challenges posed by aberrant SAAs, a rare but clinically significant condition. The findings directly inform several core competency domains in cardiovascular medicine.

#### Medical Knowledge

Clinicians should recognize that aberrant SA anatomy, defined as the SA arising from the SMA rather than the CA, is an independent risk factor for SAA formation. Patients with this variant are at increased risk of developing large, proximally located aneurysms that may rupture if left untreated.

#### Patient Care and Procedural Skills

The results highlight the importance of targeted CTA screening in patients incidentally found to have this variant, with particular attention to the proximal SA. The zs_abeSA predictive model provides a practical, CTA-based tool for individualized risk stratification, enabling clinicians to tailor surveillance intervals and consider earlier intervention in high-risk patients. This approach supports informed shared decision-making with patients regarding surveillance frequency and timing of intervention.

#### Systems-Based Practice and Practice-Based Learning

Integration of the zs_abeSA model into routine radiology and vascular surgery workflows has the potential to standardize the evaluation of patients with aberrant SA anatomy across different practice settings. The model’s reliance on standard CTA measurements facilitates adoption without requiring specialized computational expertise, promoting consistent, evidence-based care.

### Translational outlook

Our findings bridge basic hemodynamic principles with clinical practice by establishing a mechanistic link between morphological abnormalities and aneurysm formation. The integration of CFD with ML demonstrates how advanced computational techniques can be translated into user-friendly bedside tools. The web-based zs_abeSA model, requiring only standard CTA measurements, represents a low-cost, scalable solution that can be readily implemented in routine clinical workflows without specialized computational expertise. Future prospective studies are warranted to validate the model’s utility in guiding clinical decision-making and improving patient outcomes. This approach may also serve as a paradigm for studying other rare VAAs and anatomical variants, potentially extending to other vascular territories.

#### Highlights

- Aberrant SA anatomy is an independent risk factor for SAA formation.
- Aberrant SAAs are larger, more proximal, and hemodynamically distinct from common aneurysms.
- A morphology-based ML model (zs_abeSA) predicts aneurysm occurrence and location (AUROC = 0.828).
- A web-based tool derived from this model enables individualized risk stratification and clinical decision-making.

## Data Availability

The source code and model files generated during this study are available in the GitHub repository: https://github.com/RanXu1995/zs_abeSA-model. The raw imaging data are not publicly available due to patient privacy restrictions but are available from the corresponding author upon reasonable request.

## Abbreviations

SA: splenic artery
SMA: superior mesenteric artery
SAAs: splenic artery aneurysms
VAAs: visceral artery aneurysms
CFD: computational fluid dynamics
3D: three-dimensional
ML: Machine learning
WSS: wall shear stress
OSI: oscillatory shear index
CTA: computed tomography angiography

## Acknowledgments

The authors thank all participating centers and their staff for their contributions to patient enrollment and data collection. We are grateful to the Department of Radiology at each center for providing high-quality CTA images and technical support. We also thank Boea Wisdom Company (Hangzhou, China) for assistance with CFD simulations and for valuable discussions regarding the ML analysis.

## Figure titles and legends

**Central Illustration:**
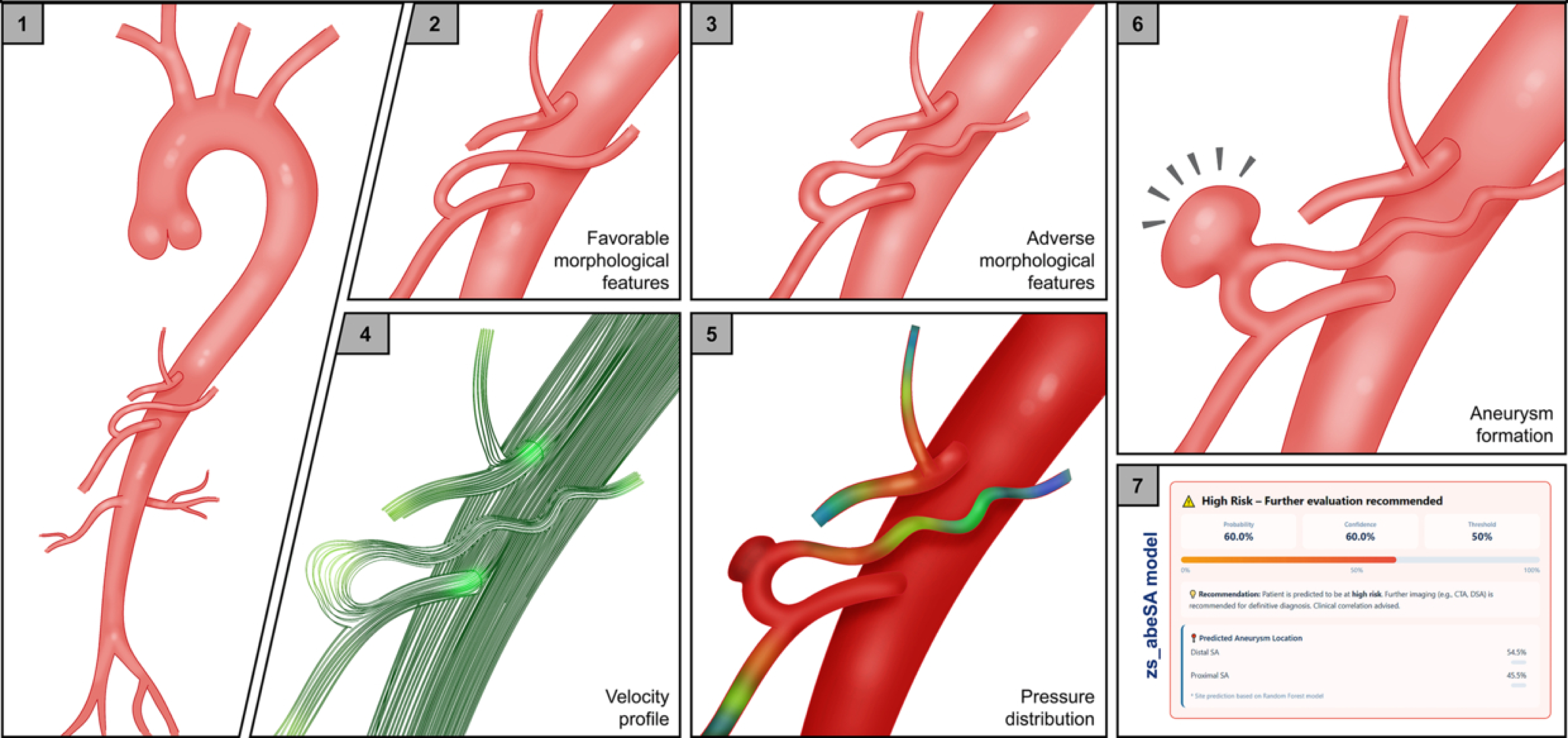
From Anatomy to Aneurysm: Morphological-Hemodynamic Coupling and Predictive Modeling in Aberrant SA. This illustration summarizes the pathogenic cascade and clinical translation of aberrant SAAs. The SA arises from the SMA rather than the CA, leading to distinct morphological alterations. These morphological features create an abnormal hemodynamic environment, particularly at the splenic artery-parent vessel junction. This hemodynamic derangement predisposes to aneurysm formation, predominantly in the proximal SA, with larger aneurysm dimensions. The zs_abeSA predictive model enables individualized risk stratification and supports clinical decision-making for screening, surveillance, and intervention strategies. CA, celiac axis. SA, splenic artery. SAA, splenic artery aneurysm. SMA, superior mesenteric artery.

